# Transmission timing and containment risk in human Andes virus outbreaks

**DOI:** 10.64898/2026.10.10.26365227

**Authors:** Nyall Jamieson, Rémy Pasco, Sam Abbott, Justin Lessler, Lauren Ancel Meyers

## Abstract

The 2026 multinational Andes virus (ANDV) outbreak linked to cruise-ship travel from Argentina triggered extensive contact tracing and quarantine. Whether intervention on this scale was necessary or sufficient depends on the risk of onward transmission, which can be difficult to estimate early in an outbreak. Using transmission-pair data from the 2018–2019 outbreak in Epuyén, Argentina, and a mechanistic model linking viral growth, symptom onset, and transmission, we estimated that 42.4% (95% CrI: 18.2–69.7%) of transmission occurs before symptom onset and that the median incubation period is 18.5 days (95% CrI: 16.5–19.5). We simulated outbreaks under an Epuyén-calibrated estimate (R = 2.12, k = 0.316) and a pooled estimate from 15 prior ANDV investigations (R = 0.24, k = 0.39), with intervention beginning after two generations, as in the cruise-associated event. Under the higher-transmission scenario, missing 10% of exposed individuals or downstream contacts resulted in a 29.9% probability of continued spread and a 20.6% probability that at least 5,000 ultimately uninfected people would be quarantined. Under lower transmission, continued spread stayed below 8%. However, the risk of onward transmission could not be estimated precisely from the first several detected cases. These findings therefore support rapid, comprehensive contact tracing and quarantine after large international exposure events despite substantial costs.

## Introduction

Andes virus (ANDV) is the only hantavirus known to be capable of person-to-person transmission, although documented transmission has been confined to a small number of localized outbreaks [1–3]. ANDV infection is associated with a case-fatality rate of approximately 36% [4], and there are currently no approved vaccines or specific antiviral therapies. Recent increases in ANDV activity in South America have heightened concern about the potential for person-to-person spread [5]. In 2026, a multinational ANDV exposure event linked to cruise-ship travel from Argentina led to 13 reported cases and 3 deaths, and prompted extensive international contact tracing and prolonged quarantine of 147 passengers and crew from 23 countries [6, 7]. The severity of ANDV disease and its potential for person-to-person spread may warrant such intensive responses, but evidence on their effectiveness and associated quarantine burden remains limited.

Evaluating containment strategies requires a detailed understanding of ANDV transmission dynamics. Decisions about the scale and duration of contact tracing and quarantine depend on incubation periods, the timing of infectiousness relative to symptom onset, and individual-level variation in transmission. Long incubation periods prolong monitoring and quarantine, transmission before symptom onset allows infected individuals to spread infection before they are recognized and isolated, and variation in transmission means that a small number of infected individuals may account for a disproportionate share of secondary infections. However, estimating these quantities from outbreak data is complicated by evolving control measures, including case detection, isolation, and quarantine, which alter transmission opportunities over time.

The 2018–2019 Epuyén outbreak provides the most detailed empirical data on person-to-person ANDV transmission. The original outbreak investigation reconstructed transmission chains, identified superspreading, and estimated that the effective reproduction number (*R*_*e*_) declined from 2.12 before implementation of isolation and quarantine to 0.96 afterward [13]. Most reported ANDV infections, by contrast, arise from zoonotic spillover and are not followed by sustained person-to-person transmission. Transmission in Epuyén concentrated at a birthday party and a wake, in a town of fewer than 2,000 people, suggesting that what distinguishes these outcomes is not where infection is acquired but the number and intensity of close contacts during the brief window around symptom onset, variation in infectiousness across individuals, or both. The relative contributions of contact and infectiousness cannot be resolved from existing investigations, which report secondary cases but not contacts that did not result in transmission. Because incubation can last several weeks, the setting an infected person occupies during the transmission window may differ substantially from the setting in which exposure occurred. Transmission potential can therefore vary among cases, locations, and generations of the same outbreak.

More recently, a latent-time analysis of the Epuyén transmission-pair data jointly estimated unobserved infection times, generation intervals, transmission timing relative to source symptom onset, time-varying reproduction numbers, and offspring dispersion from interval-censored data [12]. However, it modeled incubation and transmission timing as separate empirical delay distributions rather than as outcomes of a shared within-host process. This limited its ability to represent biological dependencies among infection, symptom onset, and infectiousness.

Here, we extend the latent-time framework by integrating a mechanistic model that jointly links infection, symptom onset, and transmission [15]. We incorporate the resulting estimates in stochastic branching-process simulations of a multinational exposure event based on the initial timing and magnitude of the 2026 cruise-associated outbreak. Given the variation in transmission observed across past ANDV events, we simulate outbreaks in both high- and low-transmission settings. The high-transmission setting uses parameters estimated from the Epuyén investigation, whereas the low-transmission setting uses pooled estimates from 15 prior ANDV investigations. We estimate the level of contact tracing and quarantine required for containment and quantify the consequences of incomplete or delayed intervention.

## Results

We fitted the within-host-informed latent-time model to 34 cases and 33 reconstructed source-recipient pairs from the 2018–2019 Epuyén outbreak in Argentina (Table 1; Figure 1). Transmission was concentrated near source symptom onset, with an estimated 42.4% of transmission events occurring before symptoms appeared (95% CrI: 18.2–69.7%). The median transmission time was 0.07 days after source symptom onset (95% CrI: (−0.20) to 0.30 days), and the central 95% of transmission events occurred from 0.85 days before to 2.02 days after source symptom onset (Figure 1A).

**Table 1:** Estimated epidemiological characteristics of person-to-person ANDV transmission. Median estimates are reported with 95% credible intervals (CrIs), reflecting uncertainty in the estimated population median. The central 95% interval summarizes the expected range across individual infections or transmission events. Transmission timing is reported relative to source symptom onset, with negative values indicating transmission before symptom onset.

| Quantity | Median estimate (95% CrI) | Central 95% interval |
| --- | --- | --- |
| Incubation period | 18.50 days (16.50–19.50) | 7.50–35.50 days |
| Latent period | 13.50 days (10.50–15.50) | 5.50–22.50 days |
| Generation interval | 22.50 days (20.50–23.50) | 12.50–38.50 days |
| Serial interval | 21.79 days (21.28–22.18) | 12.20–36.94 days |
| Transmission timing (relative to symptom onset) | 0.07 days (-0.20–0.30) | -0.85–2.02 days |

**Figure 1:**
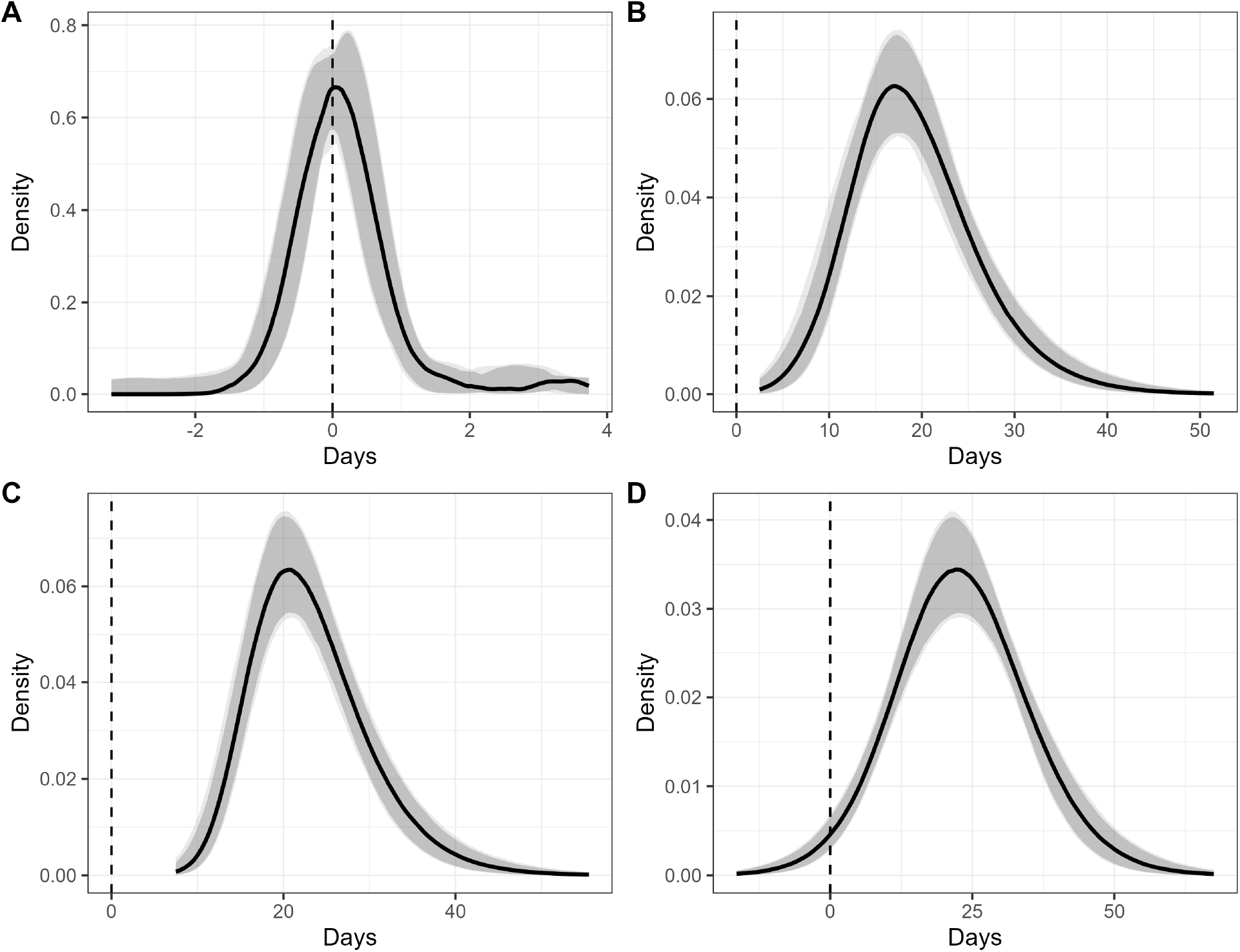
Estimated epidemiological timing distributions for person-to-person ANDV transmission. Panels show the inferred distributions of the (A) transmission timing relative to source symptom onset, (B) generation interval, (C) incubation period, and (D) serial interval. Solid curves represent posterior median densities, and shaded regions indicate the corresponding 95% credible intervals. Dashed vertical lines mark zero days. In panel A, negative values indicate transmission before source symptom onset; in panel D, negative values indicate that the recipient developed symptoms before the source case.

The median incubation period was 18.50 days (95% CrI: 16.50–19.50 days), with a central 95% distributional interval of 7.50–35.50 days (Figure 1C). An estimated 95% of infections developed symptoms by 31.50 days (95% CrI: 29.50–35.50), and 99% by 39.50 days (95% CrI: 35.50–44.50). The median generation interval was 22.50 days (95% CrI: 20.50–23.50 days) (Figure 1B), and the median serial interval was 21.79 days (95% CrI: 21.28–22.18 days) (Figure 1D). Thus, exposed individuals may require several weeks of follow-up even though onward transmission is concentrated within a narrow window around symptom onset. These estimates were broadly consistent with previous analyses of the Epuyén outbreak while providing model-based inference of latent infection times and transmission timing relative to symptom onset (Supplementary Results).

To characterize the transmission conditions documented across settings, we analyzed 15 published ANDV investigations. A hierarchical negative-binomial model estimated a pooled reproduction number of 0.24 (95% CrI: 0.12–0.48) and a shared dispersion parameter of 0.39 (95% CrI: 0.19–0.89), reflecting low average but highly variable realized transmission across this broader set of investigations. Together with the Epuyén-calibrated estimates (*R*=2.12, *k* =0.316), these results anchored an empirically informed range of ANDV transmission potential. Hereafter, we refer to simulations using the Epuyén and pooled parameterizations as the higher- and lower-transmission scenarios, respectively.

The primary analysis reproduced the timing of the 2026 cruise-associated response, with intervention beginning on simulation day 35, 26 days after symptom onset in the first detected case. We restricted the analysis to outbreaks that had reached at least two generations of transmission by then; this occurred in approximately 35% of higher-transmission simulations and 3% of lower-transmission simulations. Tracing failures had substantially greater consequences under higher transmission (Figure 2A). When tracing missed 5%, 10%, 25%, or 50% of exposed individuals or downstream contacts, post-response transmission outside quarantine occurred in 18.9%, 29.9%, 51.1%, and 70.8% of simulations, respectively. Under the lower transmission, this probability ranged from 0.7% with 5% missed to 7.8% with 50% missed. Among simulations with higher transmission and at least one additional post-response infection, missing 10% of contacts resulted in a median of 5 additional infections and a 95th percentile of 49. With 25% or 50% missed, the medians increased to 8 and 27, respectively, and the 95th percentiles to 111 and 351. With lower transmission, the median remained between one and two across tracing levels (Figure 2B).

**Figure 2:**
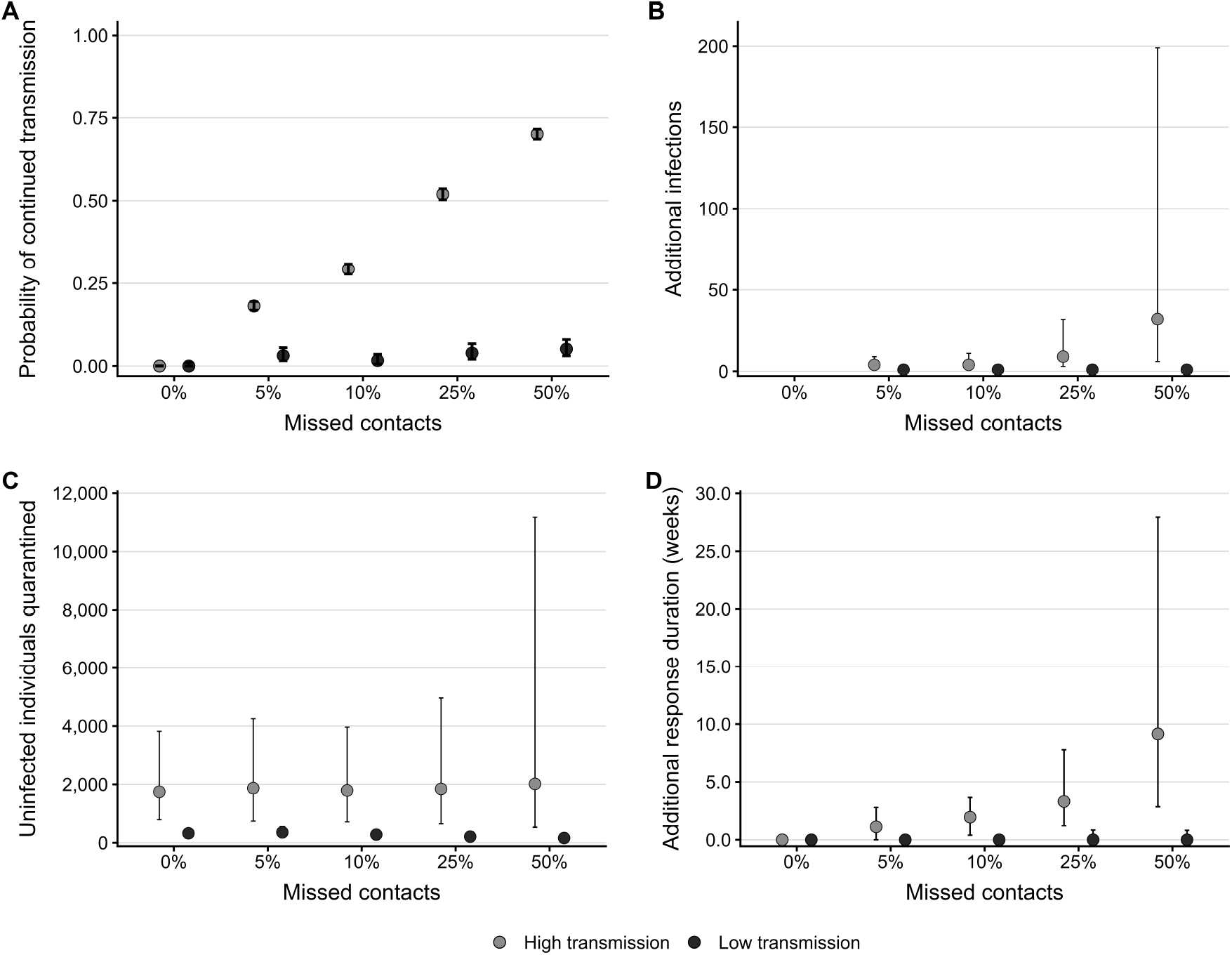
Projected outbreak and quarantine outcomes under incomplete tracing and contrasting transmission scenarios. Simulations compared an Epuyén-calibrated higher-transmission scenario and a pooled lower-transmission scenario, with intervention initiated 26 days after symptom onset of the first detected case. Simulations assume 95% case detection, a gamma-distributed delay from symptom onset to detection with mean 2 days and SD 1 day, a gamma-distributed delay from detection to isolation with mean 0.5 days and SD 0.5 days, 99% effective isolation, and a fixed 42-day quarantine. (A) Probability of at least one post-response transmission outside quarantine; points show estimated probabilities and error bars indicate exact 95% confidence intervals. (B) Additional infections conditional on at least one additional post-response infection; points show medians and error bars show interquartile ranges. (C) Downstream retrospectively uninfected contacts quarantined through post-response contact tracing, excluding the initial exposed cohort; outcomes were summarized across all conditioned simulations, with points showing medians and error bars showing interquartile ranges. (D) Additional response duration beyond the initial six-week quarantine period; outcomes were summarized across all conditioned simulations, with points showing medians and upper error bars showing the 95th percentile. Outcomes were evaluated among simulations in which the uncontrolled potential transmission tree had reached generation 2 by the response day.

Quarantine burdens were substantially lower under lower transmission (Figure 2C). With 10% of contacts missed, the median number of retrospectively uninfected downstream contacts quarantined was 313 (IQR 185–440), compared with 1,821 (IQR 720–4,161) under the higher-transmission scenario. At lower transmission, the median burden fell by 49%, from 329 with complete tracing to 167 with 50% of contacts missed, because outbreaks were small and fewer contacts were traced. At higher transmission, fewer quarantines per case were offset by larger outbreaks and more tracing events, leaving the median comparatively stable but increasing the risk of very large quarantine burdens.

Most response activity ended within the initial six-week quarantine period, but breakthrough transmission occasionally prolonged the response under the higher-transmission scenario. With 10% of contacts missed, the median additional response duration was zero months, the 95th percentile was 1.61 months, and 0.2% of simulations required at least four additional months of response activity. With 50% missed, the median increased to 1.02 months and the 95th percentile to 5.01 months. With lower transmission, additional response duration remained near zero (Figure 2D).

The multinational response to the 2026 cruise-associated outbreak was initiated after seven confirmed or suspected cases were identified. To assess the consequences of delayed intervention, we initiated contact tracing and quarantine after different numbers of cases had been reported, assuming 95% tracing completeness (Figure 3). Under higher transmission, delaying intervention from the first to the twentieth reported case increased the probability of at least 100 infections from 2.7% to 38.2% and of at least 500 infections from 2.1% to 14.0%. Median outbreak size increased from 1 to 72 infections, median quarantine burden from 2 to 500 individuals, and the probability of quarantining at least 1,000 individuals from 2.7% to 36.0%. Under lower transmission, outbreaks rarely reached the later intervention points: only 13 of 10,000 simulations reached 10 reported cases, and none reached 20. Outcomes following greater delays therefore could not be estimated precisely.

**Figure 3:**
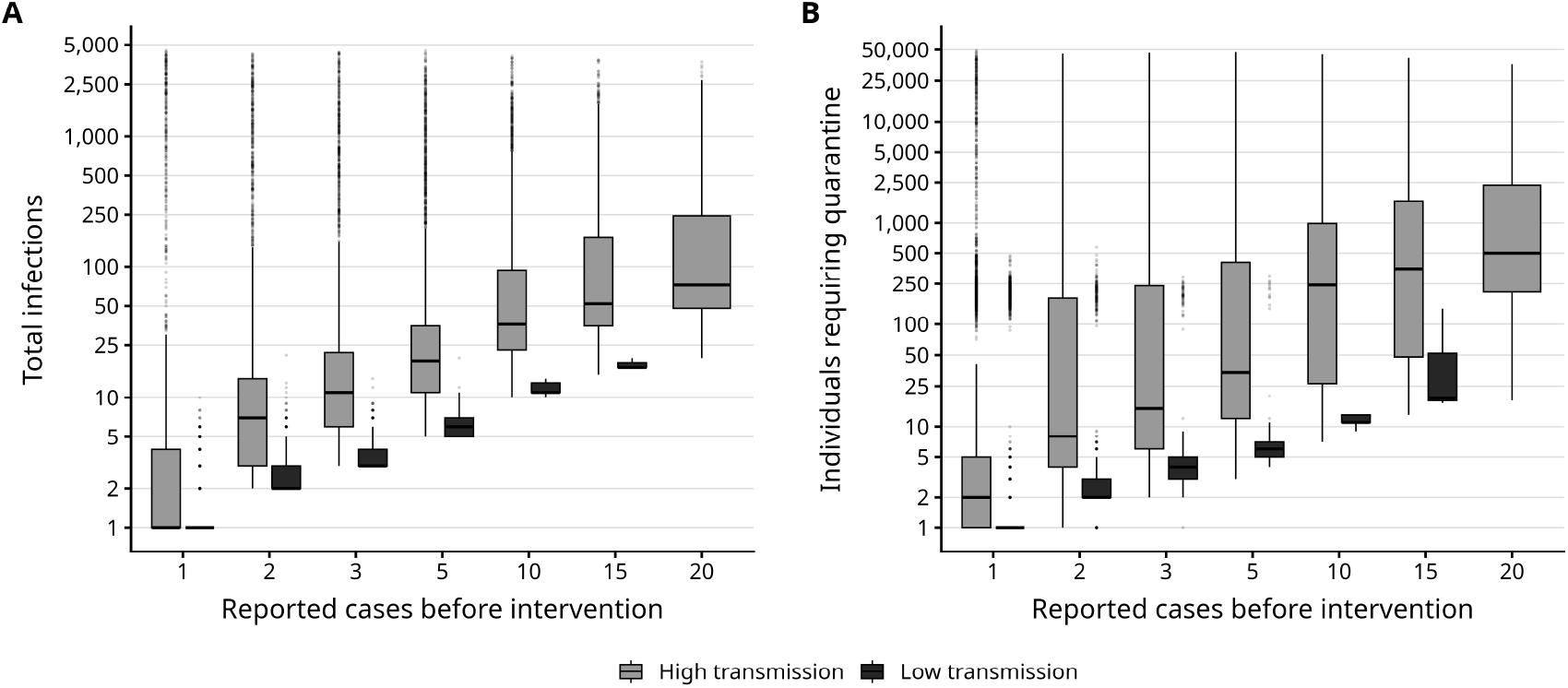
Outbreak and quarantine outcomes under delayed intervention and contrasting transmission scenarios. Simulations assumed 95% tracing completeness and varied the number of reported cases after which tracing and quarantine were initiated. Panels show the estimated distributions of (A) total infections and (B) individuals requiring quarantine under the higher-transmission (gray) and lower-transmission (black) scenarios across 10,000 stochastic simulations per transmission scenario and intervention delay. For each reported-case trigger, boxplots show the median, interquartile range, and 1.5 × interquartile-range whiskers; points represent individual simulations beyond the whiskers. Outcomes were summarized among simulations that reached the specified reported-case trigger. The y-axes are shown on a logarithmic scale to visualize the highly right-skewed outcome distributions.

Under active containment, most simulated outbreaks ended before enough cases were detected to estimate the reproduction number precisely. Estimates from the first few detected cases were highly uncertain, with broad 95% credible intervals that commonly included *R* = 1 under both transmission scenarios (Supplementary Figure 5). Among lower-transmission simulations, outbreaks that accumulated enough detected cases to inform estimation were rare realizations with unusually high transmission that resembled higher-transmission outbreaks. However, the size of the 2026 cruise-associated outbreak was much more probable under higher transmission: outbreaks of at least 13 cases occurred in 29.4% of higher-transmission simulations compared with 0.01% of lower-transmission simulations. Beyond final outbreak size, the observed timing of case accumulation was also poorly compatible with the pooled parameterization (*p*_compat_ = 0.0055). Nevertheless, this parameterization occasionally produced outbreaks of the observed magnitude.

The transmission scenarios were therefore difficult to distinguish from the first few detected cases: within-outbreak 95% credible intervals for *R* spanned 1 under both scenarios through 5 detected cases (Supplementary Figure 5). Lower-transmission outbreaks rarely reached more than 5 detected cases. This early uncertainty is compounded by the potential for transmission conditions to change as cases disperse into different settings. Delaying intervention can therefore come at a high cost, particularly if subsequent cases encounter greater opportunities for transmission.

## Discussion

The 2026 multinational ANDV exposure event linked to cruise-ship travel from Argentina prompted an extensive international contact-tracing and quarantine response. Using a mechanistic model linking viral growth, symptom onset, and transmission together with stochastic outbreak simulations, we re-estimated key transmission intervals and examined how delays in intervention or incomplete tracing affect outbreak risk and quarantine burden. We estimated that the median incubation period is 18.5 days and that transmission is concentrated within a narrow window around symptom onset, with approximately 42% occurring just before symptoms appear. Because weeks may pass between exposure and this brief transmission window, an infected person may be in a markedly different social setting by the time transmission is most likely, as when passengers dispersed from the cruise ship to 23 countries. Our simulations covered a range of transmission conditions drawn from prior ANDV investigations. Under higher transmission, modest tracing gaps or delays sharply increased both the risk of continued spread and the chance of very large quarantine burdens. Because the relevant transmission context may change across cases, places, and generations—and may not be apparent from early case counts—large international ANDV exposure events warrant rapid, comprehensive contact tracing and quarantine.

Under the Epuyén-calibrated higher-transmission scenario (*R* = 2.12, dispersion parameter *k* = 0.316), missing 10% of exposed individuals or downstream contacts resulted in a 29.9% probability of post-response transmission outside quarantine (95% CI: 28.3%–31.5%). Among simulations with at least one additional post-response infection, the median number of additional infections was 5. Across all conditioned simulations, 20.6% resulted in quarantine of at least 5,000 retrospectively uninfected downstream contacts (95% CI: 19.3%–22.1%). Risks and downstream burdens remained substantially lower under lower transmission.

A central challenge for public-health planning is that the person-to-person transmission potential of ANDV can vary markedly with context. The best-characterized outbreaks, including Epuyén and the early transmission reported during the 2026 cruise-associated outbreak, are consistent with reproduction numbers of approximately 2. Broader evidence, however, indicates that most recognized ANDV cases are isolated zoonotic infections with no identified onward transmission and that person-to-person spread has been limited to relatively few household, healthcare-associated, and community clusters in Argentina and Chile [1–3, 13, 14, 25]. These patterns are consistent with a context-dependent, highly overdispersed transmission process: many infected people transmit to no one, whereas combinations of greater infectiousness and extensive contact can produce clusters or superspreading. Our higher- and lower-transmission scenarios represent this empirically observed range rather than mutually exclusive explanations for ANDV spread.

The severity of ANDV disease typically prompts rapid intervention, while the long incubation period and concentration of transmission near symptom onset allow many exposed individuals to be quarantined before they become infectious, limiting transmission chains to a few generations. Outbreaks therefore rarely persist long enough to reveal how transmission potential varies across cases and settings. In our simulations, intervention often ended outbreaks before enough data accumulated to estimate the reproduction number precisely, and the outbreaks that did accumulate enough cases were a biased sample: the rare lower-transmission outbreaks reaching this point had unusually high realized transmission, and therefore resembled higher-transmission outbreaks.

The 2026 cruise-associated outbreak illustrates this challenge. In retrospect the scenarios are easily distinguished: an outbreak of at least 13 cases arose in 29.4% of higher-transmission simulations but 0.01% of lower-transmission simulations. This comparison requires the final outbreak size, which was unavailable when the response was initiated after seven recognized cases. Even full knowledge of transmission aboard the ship would not have established the risk after passengers dispersed, since the long incubation period allows infected people to move between settings before reaching the transmission window. Early transmission potential is therefore both poorly estimated and a poor guide to what follows. Given the consequences of failing to control an outbreak with *R* above one, tracing and isolation should proceed as if transmission is sustained.

Our within-host-informed framework builds on both the Epuyén outbreak investigation and subsequent latent-time analyses. The original Epuyén study reported incubation periods ranging from 9 to 40 days, a mean serial interval of approximately 23 days, and substantial transmission during the first few days after symptom onset, including 17 of 33 secondary infections attributed to the first day of fever and 19 of 33 occurring within the first three symptomatic days [13]. Subsequent latent-time analyses extended this descriptive epidemiology by jointly estimating latent infection times, generation intervals, transmission timing relative to symptom onset, time-varying reproduction numbers, and offspring dispersion from interval-censored transmission-pair data [12]. Our model further extends these analyses by linking infection, symptom onset, and transmission through a common biological model rather than heuristic delay distributions. Despite this more mechanistic formulation, our estimates were broadly consistent with the original descriptive investigation and subsequent latent-time analyses. They support the conclusion that transmission is concentrated near symptom onset and estimate that approximately 42% of transmission occurs before source symptom onset. The principal quantitative difference was a somewhat shorter incubation-period distribution, with a reduced upper tail relative to previous latent-time estimates. More importantly, our framework provides a mechanistic basis for these timing estimates and propagates their uncertainty directly into stochastic simulations of outbreak containment. These findings, combined with the high case-fatality rate of ANDV and the absence of approved vaccines or specific antiviral therapies, help explain why large multinational exposure events may warrant rapid, comprehensive, and potentially costly contact tracing, quarantine, and isolation measures.

Reducing the burden of large-scale quarantine will likely require improved strategies for identifying infected individuals before they can transmit infection to others. The psychological, social, and economic costs of prolonged quarantine can be substantial, particularly when most quarantined individuals ultimately prove uninfected [23, 24]. ANDV RNA has been detected by RT-PCR up to two weeks before symptom onset in some infected individuals [14], and our estimates suggest that the vast majority of transmission occurs within approximately two days of symptom onset. If serial molecular testing proves sufficiently sensitive during the incubation period, exposed individuals could potentially be managed through home monitoring with repeated testing rather than prolonged quarantine. Nonetheless, rapid case detection, isolation, and contact tracing would remain essential to contain emerging outbreaks. The 2026 multinational response therefore provides a rare opportunity to evaluate diagnostic performance during the incubation period and determine whether testing-based strategies can reduce the societal and operational costs of future responses.

Several limitations should be considered. First, the simulations represent context-dependent transmission through two fixed parameterizations and do not allow transmission conditions to change among cases or over time. The Epuyén and pooled estimates bracket transmission conditions documented in prior investigations, but neither should be interpreted as a universal reproduction number for a particular setting. Second, the pooled estimate combines studies that differed in setting, case ascertainment, control measures, and reconstruction of transmission chains. It therefore describes lower realized transmission across a heterogeneous evidence base rather than isolating the effects of contact patterns or infectiousness. Third, we make plausible but setting-nonspecific assumptions about contacts, intervention effectiveness, and other operational characteristics. Contact patterns, exposure intensity, ventilation, and other factors shaping transmission risk may differ substantially across cruise ships, households, healthcare settings, tight-knit communities, and other congregate environments. Consequently, the projected outbreak sizes and quarantine burdens should be interpreted as plausible outcomes under the modeled conditions rather than precise predictions. The modeling framework is intentionally flexible and can readily incorporate improved estimates of transmissibility, transmission timing, contact patterns, diagnostic performance, and intervention effectiveness as more data become available.

The 2026 multinational response to ANDV provides a rare opportunity to substantially improve our understanding of how ANDV transmission varies across settings and guide more effective containment strategies. The unprecedented scale of contact tracing, monitoring, and follow-up may produce data that link transmission outcomes to contact settings and substantially refine estimates of transmissibility, transmission timing, incubation-period variability, diagnostic performance, and quarantine effectiveness. The response was initiated after seven recognized cases while most exposed individuals remained aboard the ship. Had the outbreak been recognized only one or two transmission generations later, particularly after exposed individuals had dispersed internationally, containment could have required substantially larger tracing and quarantine efforts spanning multiple countries. Until additional evidence becomes available, our findings support rapid, comprehensive, and potentially costly contact tracing, quarantine, and isolation for large multinational ANDV exposure events, while highlighting the need for improved diagnostics and decision-support tools that reduce intervention burden without compromising containment.

## Methods

### Epuyén transmission-pair data

We reanalysed linked transmission-pair data from the 2018–2019 Epuyén Andes virus outbreak [13]. The analysis included 34 cases and 33 reconstructed source–recipient pairs with interval-censored exposure windows and symptom-onset information. Because exact infection and symptom-onset times were not always observed, the analysis jointly inferred latent infection times, symptom-onset times, incubation periods, generation intervals, serial intervals, and transmission timing relative to source symptom onset, building on previous latent-time analysis of the Epuyén line list [12].

### Latent infection/onset-time model

We retained the latent-time and linked-pair structure of a previous Epuyén analysis [12] but replaced its separate empirical distributions for incubation periods and transmission timing with the within-host-informed event-time model developed by [15]. By linking infection, symptom onset, and transmission through a shared within-host process, this structure regularized otherwise weakly identified event times when exposure windows were wide. It was not intended to reconstruct individual viral-load trajectories from line-list data.

For case *i* and linked transmission pair *i* → *j*, we defined the incubation period (IP), generation interval (GI), serial interval (SI), and transmission time Δ as

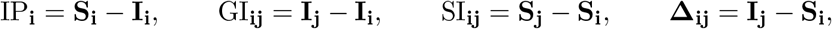

where *I*_*i*_ and *S*_*i*_ denote the infection and symptom onset time of case *i*, respectively. Negative values of Δ_*ij*_ indicate presymptomatic transmission. These quantities were defined and interpreted following standard infectious-disease timing frameworks [8–10]. Joint posterior draws were used to summarize transmission timing and parameterize the branching-process simulations while preserving dependence among the timing quantities.

The model was implemented in Stan [16]. Full model specification, priors, and posterior-derived timing quantities are provided in Supplementary Methods 1 and Supplementary Table 5.

### Transmission scenarios

We parameterized two transmission scenarios to represent the variation in realized transmission observed across prior ANDV investigations. The pooled lower-transmission scenario was estimated from published ANDV investigations across multiple settings, whereas the higher-transmission scenario was calibrated to the Epuyén outbreak.

#### Pooled lower-transmission estimate

For the lower-transmission scenario, we combined individual-level offspring counts and aggregate numbers of primary and secondary cases from 15 published ANDV outbreak and contact-investigation studies [2, 13, 14, 26–37]. For studies reporting identifiable transmission chains or explicit index–secondary case relationships, we derived the number of secondary infections attributed to each primary case directly from the original outbreak reports. For studies without individual transmission chains, we extracted the numbers of primary cases at risk of onward transmission and attributed secondary cases from the systematic-review tables. The complete individual offspring vectors and study-level aggregate counts used in the analysis are provided in the accompanying data file. Table 2 summarizes the data used to estimate the reproduction number *R* and dispersion parameter *k*.

**Table 2:**
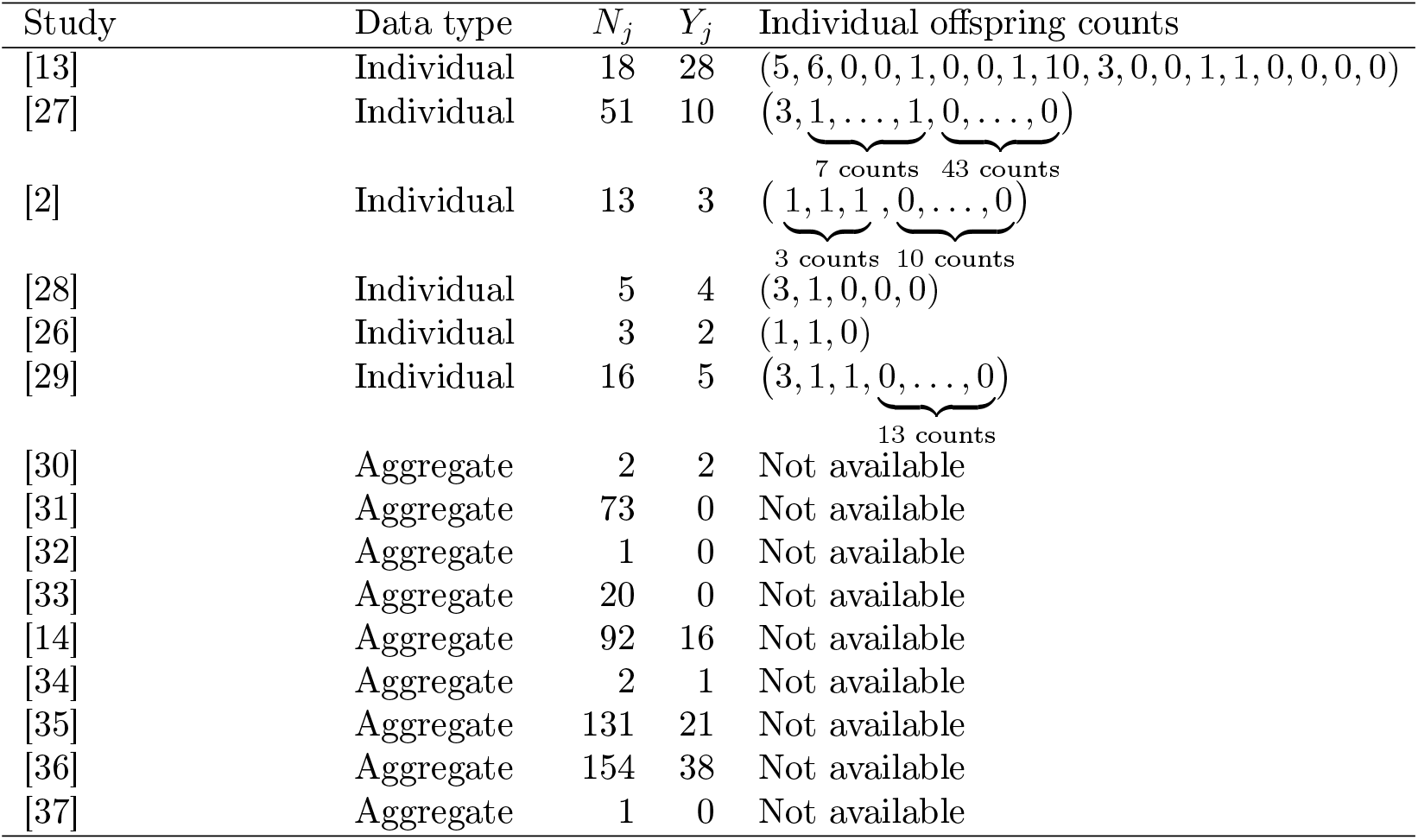
Data used in the hierarchical negative-binomial transmission model. For individual-level studies, *N*_*j*_ is the number of cases represented in the offspring vector and *Y*_*j*_ is the sum of the offspring counts. These data were reconstructed from reported transmission chains or explicit index–secondary case relationships. For aggregate studies, *N*_*j*_ is the number of primary cases at risk of onward transmission and *Y*_*j*_ is the number of attributed secondary cases. Individual offspring counts were unavailable for these studies.

Study-specific reproduction numbers were modelled hierarchically on the log scale,

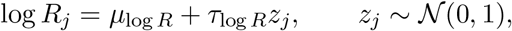

with a shared negative-binomial offspring-dispersion parameter *k*. For individual-level studies, each offspring count was modelled with mean *R*_*j*_, dispersion *k*, and variance 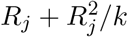. For aggregate studies, the total number of secondary cases among *N*_*j*_ primary cases was modelled using a negative-binomial distribution with mean *N*_*j*_*R*_*j*_ and dispersion *N*_*j*_*k*. We defined the pooled reproduction number as exp(*µ*_log*R*_), which corresponds to the median of the study-specific reproduction-number distribution. We used the priors

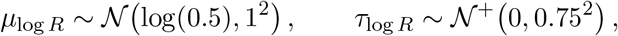

and

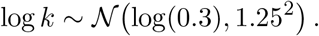

The model was fitted in Stan using four chains, 2,000 warm-up iterations and 4,000 retained iterations per chain. Sampling used `adapt_delta = 0.99, max_treedepth = 15`, and random seed 2026.

The analysis yielded the following offspring distribution, used for the lower-transmission simulations:

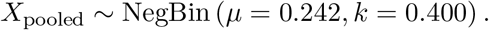

#### Epuyén-calibrated higher-transmission estimate

For the higher-transmission scenario, we fixed the negative-binomial mean at the pre-intervention effective reproduction number reported for Epuyén *R* = 2.12, and estimated the dispersion parameter by maximum likelihood from the observed Epuyén offspring counts provided in the first row of Table 2. This yielded a dispersion of 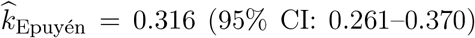. Thus, the offspring distribution used for the higher-transmission simulations was

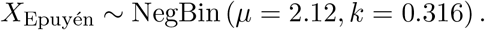

### Compatibility of the cruise-associated outbreak with transmission scenarios

To assess whether the observed 2026 cruise-associated outbreak was compatible with the two transmission scenarios, we compared its symptom-onset trajectory with stochastic outbreaks generated under each parameterization. The observed epidemic curve comprised 13 reported cases and was aligned to the date of the first symptom onset and aggregated into weekly incidence. For each transmission scenario, we simulated 10,000 outbreaks using the corresponding negative-binomial offspring distribution and the estimated generation-interval and incubation-period distributions. Simulated onset trajectories were likewise aligned to their first symptom onset and aggregated to weekly incidence. We quantified compatibility using the Mahalanobis distance between the observed weekly epidemic curve and the mean simulated trajectory, accounting for the covariance in incidence among weeks. An empirical compatibility probability was calculated as the proportion of simulated outbreaks with a Mahalanobis distance at least as large as that of the observed outbreak. As an additional measure of compatibility with the observed outbreak magnitude, we calculated the proportion of simulations that generated at least 13 cases. Further implementation details are provided in the Supplementary Methods.

### Branching-process model of ANDV transmission and containment

We developed a stochastic individual-level branching-process model to evaluate person-to-person ANDV transmission after initiation of a public-health response to an international exposure event under the two transmission scenarios defined above (Figure 4). Following established early-outbreak containment models, we represented case detection, isolation, contact tracing, and quarantine as individual-level events and allowed some infections to remain undetected [17–20].

**Figure 4:**
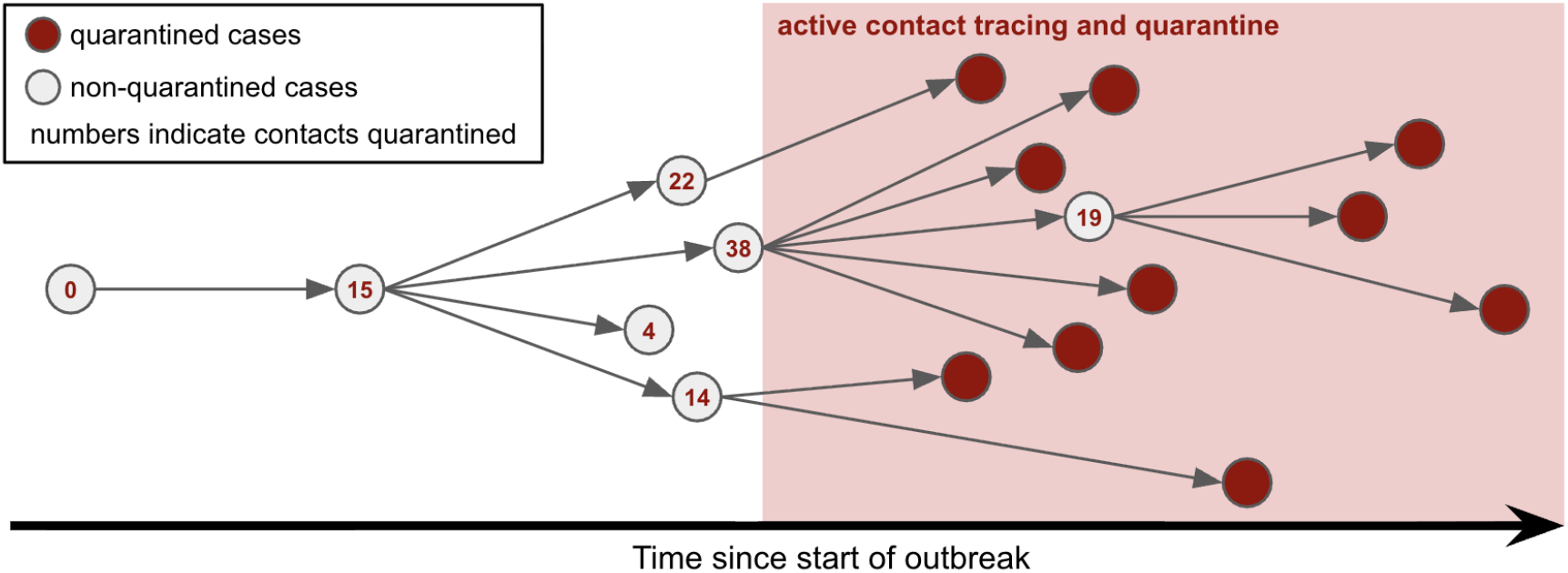
Schematic of transmission, contact tracing, and quarantine in the outbreak simulation model. Circles represent infected individuals and arrows represent transmission events, with circles positioned according to symptom-onset date. The red shaded region indicates the period of public-health response. Red circles indicate infected individuals quarantined through the initial response or subsequent contact tracing; open circles indicate infected individuals who were not quarantined before onward transmission. Numbers indicate contacts quarantined following case detection. The figure illustrates how incomplete identification of exposed individuals can allow continued transmission and generate large downstream outbreak and quarantine burdens.

We ran 10,000 simulations per transmission and intervention scenario with computational limits listed in Table 3. No simulation reached either limit; therefore, all simulated outbreaks ended before the computational boundaries, and no truncation-related exclusions or special classifications were required.

**Table 3:**
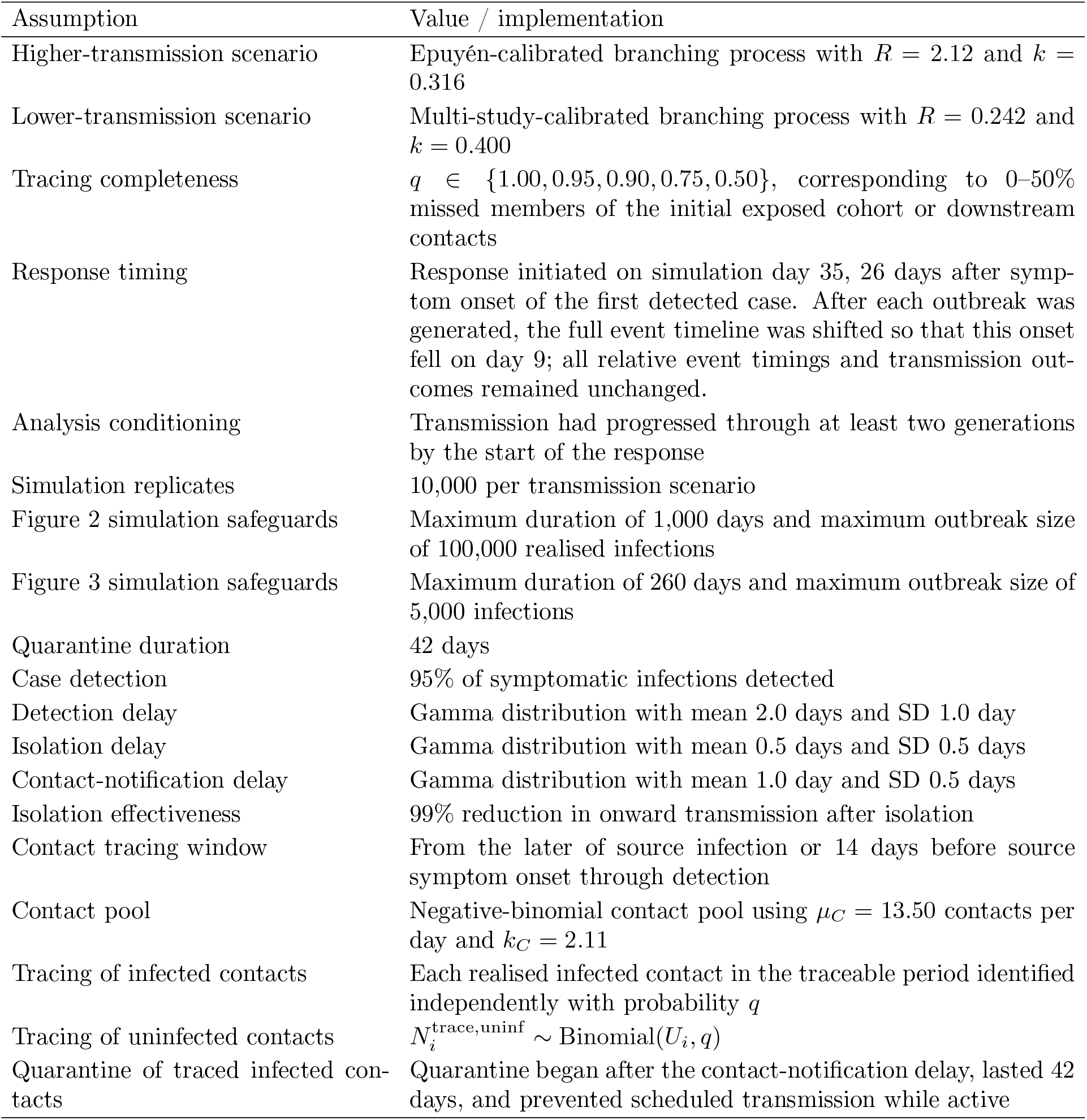
Key modeling assumptions. Full parameter definitions, derived quantities, prior settings, and implementation details are provided in Supplementary Methods 1–2 and Supplementary Tables 4–5.

Each simulation began with one introduced infection and proceeded as a chronological event-driven branching process. For each realised infection, potential transmission events were scheduled using the scenario-specific negative-binomial offspring distribution, with generation intervals and recipient incubation periods sampled jointly from the posterior timing draws. Symptom onset, case detection, isolation, contact tracing, quarantine, and transmission events were then processed in chronological order.

Before response initiation, transmission proceeded without intervention. After the response began, each scheduled transmission was evaluated according to the quarantine and isolation status of the source at the time of transmission. Transmissions prevented by quarantine or isolation did not generate recipient infections or downstream transmission branches. Each simulation ended naturally when no future events remained, subject only to pre-specified computational safeguards.

We then evaluated whether a fixed public-health quarantine response captured infected individuals in the known exposed cohort and prevented subsequent transmission.

### ANDV outbreak response scenario

The mitigation model is based on the international response to the 2026 cruise-associated outbreak. We assumed that public-health authorities identify an initial cohort of exposed individuals. Infected individuals with infection times on or before the start of the response were considered members of this cohort and were eligible for quarantine.

The response began on simulation day 35 in every simulation. After each stochastic outbreak trajectory was generated, we shifted its entire timeline so that symptom onset of the first detected case fell on day 9. This place the response initiation 26 days after symptom onset while preserving the relative timing of infection, symptom onset, detection, isolation, and transmission events. Because many introductions extinguished spontaneously, the primary analysis included only realised outbreaks that had progressed through at least two generations of transmission by response initiation, thereby focusing on established transmission chains rather than self-limiting introductions.

At the start of the response, infected members of the known exposed cohort were quarantined independently with probability *q*. The same *q* was used as the probability of successfully tracing each later infected or uninfected contact. This was a simplifying assumption that imposed equal completeness for initial cohort identification and downstream contact tracing. We evaluated *q* ∈ {1.00, 0.95, 0.90, 0.75, 0.50}, corresponding to 0%, 5%, 10%, 25%, and 50% missed exposed individuals or later contacts.

The intervention assumptions included a fixed 42-day quarantine, 95% case detection, a gamma-distributed onset-to-detection delay with mean 2.0 days and SD 1.0 day, a gamma-distributed detection-to-isolation delay with mean 0.5 days and SD 0.5 days, and a 99% reduction in onward transmission after isolation. These delay distributions were operational assumptions rather than estimates from ANDV data.

### Contact tracing and quarantine burden

For detected cases after response initiation not already under quarantine, we reconstructed a traceable contact pool to estimate downstream quarantine burden. The traceable period ran from the later of the source case’s infection time and 14 days before symptom onset until detection. If *d*_*i*_ denotes this duration, the total contact pool was drawn as

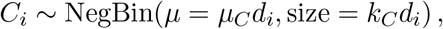

with *µ*_*C*_ = 13.50 contacts per day, *k*_*C*_ = 2.11, and a contact multiplier of 1, based on social-contact survey data [21, 22]. If 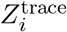 denotes the number of realised infected contacts in this period, we set

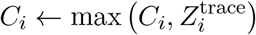

to ensure that the reconstructed contact pool included all realised infected contacts. The number of retrospectively uninfected contacts in the pool was then

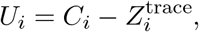

Each infected contact within the traceable period was traced independently with probability *q*. The number of traced uninfected contacts was sampled as

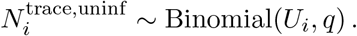

Traced uninfected contacts were added to the downstream uninfected-quarantine burden. For each successfully traced infected contact, quarantine began after a gamma-distributed notification delay with mean 1.0 day and SD 0.5 days and lasted for 42 days. Detection, tracing, quarantine, and potential-transmission events were processed chronologically, allowing quarantine to prevent subsequently scheduled transmission events.

Primary outcomes were the probability of at least one post-response transmission occurring outside quarantine, the number of additional post-response infections conditional on at least one such infection, the number of retrospectively uninfected downstream contacts quarantined through post-response contact tracing, and additional response duration beyond the initial six-week quarantine period. The downstream quarantine-burden outcome excluded uninfected members of the initial exposed cohort. Quarantine burden and additional response duration were summarized across all conditioned simulations, including simulations with no additional post-response infection. Total response duration was first calculated from response initiation until completion of all subsequent simulated transmission, detection, tracing, and quarantine activity. Additional response duration was then defined as

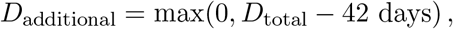

and converted to months using 30.4375 days per month.

## Data Availability

All analysis code, Stan model files, simulation scripts, and data-processing scripts needed to reproduce the analyses are available at: \url{https://github.com/epiENGAGE/Hantavirus-paper}.

