## Supplementary materials for "Transmission timing and containment risk in human Andes virus outbreaks"

#### Code availability

All analysis code, Stan model files, simulation scripts, and data-processing scripts needed to reproduce the analyses are available at: `https://github.com/epiENGAGE/Hantavirus-paper`.

#### Supplementary Methods and model assumptions

This section provides the full assumptions, parameter values, and implementation details for the linked modelling components used in the main analysis. Supplementary Table 4 describes the two-pass stochastic branching-process model used for cruise-response scenario analyses under the Epuyén-calibrated higher-transmission and pooled lower-transmission scenarios. This model first generated an uncontrolled potential transmission tree and then replayed potential transmissions after imposing a fixed public-health response. For each potential transmission tree, the outbreak calendar was aligned so that simulation day 35 occurred 26 days after symptom onset of the first detected case. Supplementary Table 5 describes the latent infection/onset-time model used to estimate infection times, symptom-onset times, incubation periods, generation intervals, serial intervals, and transmission timing relative to source symptom onset. Supplementary Methods 1–2 provide the mathematical and algorithmic details of the latent infection/onset-time model and the stochastic branching-process contact-tracing model.

**Table 4: Branching-process model assumptions and parameter values for the cruise-response analyses under the higher and lower-transmission scenarios.** Disease-timing quantities were sampled from posterior distributions estimated by the latent infection/onset-time model. The cruise-response model used a two-pass simulation design: an uncontrolled potential transmission tree was first generated, and potential transmissions were then replayed in chronological order after imposing a fixed public-health response. For each potential transmission tree, the outbreak calendar was aligned so that simulation day 35 occurred 26 days after symptom onset of the first detected case. The response represented capture and quarantine of the known exposed cohort, with downstream contact tracing around detected post-response cases used to estimate subsequent quarantine burden. The higher-transmission parameterization was used as a high-consequence scenario and should not be interpreted as an estimate of typical endemic ANDV transmissibility.

| Component | Variable | Value / distribution |
|---|---|---|
| Infector | $i$ | Simulated source case in the branching process. |
| Infectee | $j$ | Simulated recipient case generated by source $i$. |
| Initial infections | $n_0$ | 1 introduced infection per simulation. |
| Simulation replicates | $N_{\text{sim}}$ | 10,000 simulations per scenario. |

*Continued on next page*

| Component | Variable | Value / distribution |
|---|---|---|
| Maximum duration | $t_{\max}$ | 1,000 days for the primary fixed day-35 cruise-response analysis shown in Figure 2; 260 days for the reported-case-trigger delayed-intervention analysis shown in Figure 3. |
| Maximum simulated cases | $C_{\max}$ | 100,000 realised infections for the primary fixed day-35 cruise-response analysis shown in Figure 2; 5,000 infections for the reported-case-trigger delayed-intervention analysis shown in Figure 3. |
| Transmission generation | $g_i$ | Index case assigned generation 0; each retained offspring assigned $g_j = g_i + 1$. |
| Current generation | $g_{\text{current}}$ | $g_{\text{current}} = 2$, representing the current generation-2 quarantine/response point. |
| Current-generation conditioning | – | Primary cruise-response analyses were conditioned on the uncontrolled potential transmission tree reaching generation 2 by the fixed day-35 response. This conditioning represents a high-risk exposure scenario requiring public-health response, not the average outcome of an arbitrary ANDV spillover infection. |
| Tracing-start generation | $g_{\text{start}}$ | Sensitivity analysis only: $g_{\text{start}} \in \{0, 1, 2, 3, 4\}$. For each replicate, the response time was defined as the earliest infection time among cases in generation $g_{\text{start}}$. Before this response time, transmission proceeded without intervention. At the response time, infected individuals already present in the simulated outbreak were eligible for quarantine independently with probability $q$, regardless of transmission generation. This generation-start analysis was separate from the primary fixed day-35 cruise-response analysis and from the reported-case-trigger delayed-intervention analysis shown in Figure 3. |
| Tracing completeness scenarios | $q$ | Primary scenarios used $q \in \{1.00, 0.95, 0.90, 0.75, 0.50\}$. |
| Response day | – | Simulation day 35. After each outbreak was generated, the full event timeline was shifted so that symptom onset of the first detected case occurred on day 9, placing response initiation 26 days after that symptom onset. |
| Exposed cohort size | – | 100 individuals in the simulated initial exposed cohort. This is a modelling assumption rather than the observed cruise-ship cohort size; uninfected members of this initial cohort were excluded from the downstream uninfected-quarantine burden reported in Figure 2C. |
| Higher-transmission potential secondary infections | $Z_i^{\text{pot}}$ | NegBin($\mu = 2.12,\ k = 0.316$) for the Epuyén-higher-transmission scenario. |
| Lower-transmission potential secondary infections | $Z_i^{\text{pot}}$ | NegBin($\mu = 0.242,\ k = 0.400$) for the pooled lower-transmission scenario. |
| Infector infection time | $I_i$ | Index infection time set to 0; descendant infection times assigned recursively using sampled generation intervals. |

*Continued on next page*

| **Component** | **Variable** | **Value / distribution** |
|---|---|---|
| Symptom-onset time | $S_i$ | Calculated as $S_i = I_i + IP_i$. |
| Incubation period | $IP_i = S_i - I_i$ | Sampled from posterior incubation-period distribution exported by the latent infection/onset-time model. |
| Proposed recipient infection time | $I_j$ | Assigned as $I_j = I_i + GI_{ij}$. |
| Generation interval | $GI_{ij} = I_j - I_i$ | Sampled from posterior generation-interval distribution exported by the latent infection/onset-time model. |
| Transmission timing relative to source symptom onset | $\Delta_{ij} = I_j - S_i$ | Derived posterior quantity used to summarize whether transmission occurred before or after source symptom onset. |
| Serial interval | $SI_{ij} = S_j - S_i$ | Derived from simulated infection times and posterior incubation-period draws. |
| Case detection | $D_i$ | Bernoulli($p_{\text{det}}$), with $p_{\text{det}} = 0.95$. |
| Detection delay | $\delta_i$ | Gamma distribution with mean 2.0 days and SD 1.0 day; operational response assumption, not estimated from Andes-virus-specific data. |
| Detection time | $T_i^{\text{det}} = S_i + \delta_i$ | Calculated from symptom-onset time and detection delay. |
| Isolation delay | $\epsilon_i$ | Gamma distribution with mean 0.5 days and SD 0.5 days; operational response assumption, not estimated from Andes-virus-specific data. |
| Isolation time | $T_i^{\text{iso}} = T_i^{\text{det}} + \epsilon_i$ | Calculated from detection time and isolation delay. |
| Isolation effectiveness | $e_{\text{iso}}$ | 0.99 once isolation is active. |
| Delayed-start intervention rule | – | Before the scenario-specific response time, isolation and quarantine did not reduce transmission. At the response time, infected individuals already present in the simulated outbreak were eligible for quarantine independently with probability $q$, regardless of generation. After the response time, active quarantine and isolation could reduce onward transmission, and newly infected retained cases could subsequently enter quarantine according to the tracing-completeness assumption. |
| Residual transmission after isolation | $r_i^{\text{iso}}(t)$ | $r_i^{\text{iso}}(t) = \begin{cases} 1, & t < t_{\text{response},r}(g_{\text{start}}), \\ 1, & t \geq t_{\text{response},r}(g_{\text{start}}) \text{ and } t < T_i^{\text{iso}}, \\ 1 - e_{\text{iso}}, & t \geq t_{\text{response},r}(g_{\text{start}}) \text{ and } t \geq T_i^{\text{iso}}. \end{cases}$ |
| Backward tracing window | $b$ | 14 days. |
| Traceable-period start | $T_i^{\text{trace,start}}$ | $T_i^{\text{trace,start}} = \max(S_i - b, I_i)$. |
| Traceable-period end | $T_i^{\text{trace,end}}$ | $T_i^{\text{trace,end}} = T_i^{\text{det}}$. |
| Traceable contact duration | $d_i$ | $d_i = \max(0, T_i^{\text{trace,end}} - T_i^{\text{trace,start}})$. |
| Distribution of daily contacts | – | NegBin($\mu_C, k_C$). |
| Daily contact rate | $\mu_C$ | 13.50 contacts per day. |
| Contact-count dispersion | $k_C$ | 2.11. |
| Contact multiplier | – | 1.0. |

*Continued on next page*

| Component | Variable | Value / distribution |
|---|---|---|
| Minimum contact-pool constraint | $C_i$ | Total contact count constrained to be at least the number of realised infected contacts in the traceable period. |
| Infected contacts in traceable pool | $Z_i^{\text{trace}}$ | Realised infected offspring of source $i$ whose infection times fall within $[T_i^{\text{trace,start}}, T_i^{\text{trace,end}}]$. |
| Uninfected contacts in traceable pool | $U_i$ | $U_i = C_i - Z_i^{\text{trace}}$. |
| Traced infected contacts | $B_{ij}^{\text{trace}}$ | Each realised infected contact $j$ whose infection time fell within the traceable period was identified independently with probability $q$: $$B_{ij}^{\text{trace}} \sim \text{Bernoulli}(q).$$ |
| Traced uninfected contacts | $N_i^{\text{trace,uninf}}$ | If $$U_i = C_i - Z_i^{\text{trace}},$$ then $$N_i^{\text{trace,uninf}} \sim \text{Binomial}(U_i, q).$$ |
| Notification delay | $\omega_{ij}$ | Gamma distribution with mean 1.0 day and SD 0.5 days. |
| Quarantine start | $T_j^Q$ | $T_j^Q = T_i^{\text{det}} + \omega_{ij}$ for traced infected contact $j$ identified from source $i$. |
| Quarantine duration | – | 42 days. |
| Quarantine end | $T_j^{Q,\text{end}}$ | $T_j^{Q,\text{end}} = T_j^Q + 42$. |
| Quarantine adherence | – | 1.0. |
| Quarantine effectiveness | $e_Q$ | 1.0. |
| Residual transmission during quarantine | $r_j^Q(t)$ | 1 outside quarantine; $1 - e_Q$ during quarantine. Quarantine effects applied only to contacts who were traced and quarantined. |
| Realised transmission probability | $r_i(t)$ | Product of residual source isolation and source quarantine probabilities at the scheduled transmission time, subject to the delayed-start intervention rule. |
| Retained transmission | – | Potential transmission retained with probability $r_i(t)$. |
| Prevented transmission | – | Potential transmission counted as prevented if not retained because of isolation or quarantine. |
| Infected contacts traced before symptoms | – | Traced infected contacts with quarantine start before or at their symptom-onset time. |
| Additional infections after current generation | $A_{\text{inf}}$ | Number of simulated cases with $g_i > g_{\text{current}}$. |
| Any additional cases after current generation | $\widehat{P}(A_{\text{inf}} > 0)$ | Proportion of simulations, conditional on reaching generation 2, with at least one additional case after $g_{\text{current}} = 2$. |
| Additional quarantined infected contacts | $A_{\text{Q,inf}}$ | Number of infected cases after generation 2 who were traced and quarantined. |
| Additional quarantined uninfected contacts | $A_{\text{Q,uninf}}$ | Number of retrospectively uninfected contacts quarantined from tracing activity at or after the current generation-2 response point. |

*Continued on next page*

| Component | Variable | Value / distribution |
|---|---|---|
| Additional total quarantine burden | $A_{\mathrm{Q,total}}$ | $A_{\mathrm{Q,total}} = A_{\mathrm{Q,inf}} + A_{\mathrm{Q,uninf}}$. |
| Total response duration | $D_{\mathrm{total}}$ | Time from the fixed day-35 response until the end of all subsequent simulated transmission, detection, tracing, and quarantine activity. |
| Additional response duration | $D_{\mathrm{additional}}$ | Duration beyond the initial 42-day quarantine period: $$D_{\mathrm{additional}} = \max\{0,\, D_{\mathrm{total}} - 42 \text{ days}\}.$$ This outcome was converted to months and used in Figure 2D. |
| Delayed-start outcomes | – | For each $g_{\mathrm{start}}$, recorded probability of any additional cases after generation 2, 95th percentile of total infections, 95th percentile of additional quarantine burden, and 95th percentile of additional response duration. |
| Recorded outcomes | – | Tracing-start and current generations, whether each was reached, scenario-specific response time, total potential and retained infections, number and occurrence of additional cases after generation 2, post-response infections and transmissions, additional quarantine burden, transmissions prevented by quarantine or isolation, and response duration after the scenario-specific response point. |

**Table 5: Latent infection/onset-time model assumptions, fixed settings, derived quantities, and prior distributions.** Rows labelled as priors give the Bayesian prior distributions used in model fitting, including implicit uniform priors induced by the constrained latent-time parameterization. The latent timing model estimated infection and symptom-onset times using a first-event symptom-onset model for incubation periods and a normalized observed-pair transmission-intensity model for linked-pair generation timing. Posterior incubation-period, observed-pair generation-interval, serial-interval, and transmission-timing distributions were exported for use in the branching-process simulations.

| Component | Quantity | Value / distribution |
|---|---|---|
| Latent onset time | $T_{\mathrm{onset},i}$ | Estimated within case-specific onset interval. |
| Latent infection time | $T_{\mathrm{inf},i}$ | Estimated within case-specific exposure interval. |
| Latent onset-time prior | $T_{\mathrm{onset},i}$ | Implicit uniform prior within the case-specific onset interval. Equivalently, the Stan parameterization uses $u_{\mathrm{onset},i} \sim \mathrm{Uniform}(0,1)$ and maps this to $T_{\mathrm{onset},i} = L_i^{\mathrm{onset}} + u_{\mathrm{onset},i}(U_i^{\mathrm{onset}} - L_i^{\mathrm{onset}})$. |
| Latent infection-time prior | $T_{\mathrm{inf},i}$ | Implicit uniform prior within the case-specific exposure interval. Equivalently, the Stan parameterization uses $u_{\mathrm{inf},i} \sim \mathrm{Uniform}(0,1)$ and maps this to $T_{\mathrm{inf},i} = L_i^{\mathrm{exp}} + u_{\mathrm{inf},i}(U_i^{\mathrm{exp}} - L_i^{\mathrm{exp}})$. |
| Incubation period | $IP_i$ | $IP_i = T_{\mathrm{onset},i} - T_{\mathrm{inf},i}$. |
| Generation interval | $GI_{ij}$ | $GI_{ij} = T_{\mathrm{inf},j} - T_{\mathrm{inf},i}$ for linked transmission pair $i \to j$. |

*Continued on next page*

| **Component** | **Quantity** | **Value / distribution** |
|---|---|---|
| Serial interval | $SI_{ij}$ | $SI_{ij} = T_{\text{onset},j} - T_{\text{onset},i}$. |
| Transmission timing relative to source symptom onset | $\Delta_{ij}$ | $\Delta_{ij} = T_{\text{inf},j} - T_{\text{onset},i}$. |
| Index-case infection window | `index_pre_onset_max` | 80 days before onset. |
| Sourced-case fallback infection-time window | – | Used only when exposure-window data are missing. The latent infection time was allowed to vary from source symptom onset −40 days to source symptom onset +80 days; this broad window is a permissive missing-data fallback rather than a known exposure interval. |
| Multiple-source policy | `multi_source_policy` | First listed source used when multiple sources are listed. |
| Within-host-informed trajectory | $V(t)$ | Rise-and-fall relative pathogen-burden trajectory: $V(t) = A\exp(rt)\{1 + \frac{r}{d}\exp[\kappa(t-m)]\}^{-(r+d)/\kappa}$. |
| Early growth parameter | $r$ | Controls early increase in the within-host-informed trajectory; prior specified on the log scale: $\log r \sim \text{Normal}(\log(0.65), 0.25)$. |
| Late decline parameter | $d$ | Controls late decrease in the within-host-informed trajectory; prior specified on the log scale: $\log d \sim \text{Normal}(\log(0.13), 0.25)$. |
| Peak-timing parameter | $m$ | Controls peak timing of the within-host-informed trajectory; prior: $m \sim \text{Normal}(14.0, 2.0)$. |
| Transition-sharpness parameter | $\kappa$ | Controls sharpness of the transition from growth to decline; prior specified on the log scale: $\log \kappa \sim \text{Normal}(\log(0.35), 0.50)$. |
| Arbitrary scale | $A$ | Relative scale only; absolute pathogen-load scale not identified from line-list data. |
| Symptom-onset hazard | $h_{\text{sym}}(t)$ | $h_{\text{sym}}(t) = \lambda_{\text{sym}}V(t)$; prior specified on the log scale: $\log \lambda_{\text{sym}} \sim \text{Normal}(\log(0.03), 0.35)$. |
| Incubation-period density | $f_{IP}(t)$ | First-event density implied by $h_{\text{sym}}(t)$ and normalized over finite support. |
| Observed-pair transmission intensity | $w_{\text{trans}}(t)$ | $w_{\text{trans}}(t) \propto V(t)$, optionally multiplied by a soft transmission-timing gate. This is a normalized intensity for realized linked transmission pairs, not a first-event transmission hazard. |
| Observed-pair generation-interval density | $f_{GI}^{\text{obs}}(t)$ | Normalized density proportional to the observed-pair transmission intensity over finite support. This describes the timing of realized linked transmission pairs, not the waiting time to first successful transmission. |
| Integration support | $t_{\max}$ | 120 days. |
| Numerical integration intervals | $K_{\text{int}}$ | 240. |
| Numerical integration step | $\Delta t_{\text{int}}$ | 0.5 days. |
| Posterior PMF export grid | `grid_day` | 0–119 days. |
| Posterior PMF grid width | $\Delta t_{\text{grid}}$ | 1.0 day. |
| Transmission-timing gate | `use_transmission_gate` | 0 in the baseline model. If enabled, the gate acts as a soft modifier of observed-pair transmission intensity; it is not a first-event latent-period or infectiousness-onset model. |
| Transmission gate onset prior | `gate_onset` | Normal prior with mean 15.0 days and SD 7.0 days; only used when transmission gate is enabled. |
| Transmission gate slope prior | log(gate_slope) | Normal prior with mean log(2.0) and SD 0.5; only used when transmission gate is enabled. |

*Continued on next page*

| Component | Quantity | Value / distribution |
|---|---|---|
| Delta mean prior | `use_delta_mean_prior` | Enabled in baseline model. |
| Mean delta prior | $\Delta_{\text{mean}}$ | Normal prior with mean −1.0 days and SD 1.5 days. |
| Control-selection correction | `use_control_selection` | Enabled in baseline model. |
| Control date | – | 2018-12-31. |
| Post-control source indicator | – | Source cases with onset date on or after the control date are treated as post-control sources. |
| Post-control isolation delay | – | 2.0 days. |
| Residual contact probability after control | – | 0.05. |
| Control-selection weight | – | Transmission events after the isolation-delay cutoff are downweighted by the residual contact probability. |
| Control-selection normalizer | – | Step-function normalizer based on the normalized observed-pair transmission-intensity CDF at source incubation plus isolation delay. |
| Observed offspring counts | $Z_i^{\text{obs}}$ | Negative binomial with mean $R_i$ and dispersion $k_{\text{NB}}$ |
| Time-varying reproduction number | $R_i$ | Evaluated at source symptom onset time using linearly interpolated log-$R$ knots. |
| Reproduction-number knots | `knot_day` | Weekly knots spanning onset times. |
| Random walk for log reproduction number | $\log R_b$ | $\log R_1 = \log R_{\text{init}}$ and $\log R_b = \log R_{b-1} + \sigma_{\text{rw}}\epsilon_b$ for $b > 1$, with $\epsilon_b \sim \text{Normal}(0, 1)$, $\log R_{\text{init}} \sim \text{Normal}(\log(1.5), 1.0)$, and $\sigma_{\text{rw}} \sim \text{Normal}(0, 0.5)$ constrained positive. |
| Offspring dispersion | $k_{\text{NB}}$ | $k_{\text{NB}} = \text{inv\_square}(\phi^{-1/2})$, with $\phi^{-1/2} \sim \text{Normal}(0, 1.0)$ constrained positive. |
| Transmission-intensity scale | $\lambda_{\text{inf}}$ | Normal(0, 1.0), constrained positive controls the timescale of the first-event infectiousness component used in the observed-pair generation-interval model; prior specified on the log scale: $\log \lambda_{\text{inf}} \sim \text{Normal}(\log(0.04), 0.35)$. |
| Prior predictive sampling | – | 4 chains, 500 warmup iterations, 500 sampling iterations. |
| Posterior sampling | – | 4 chains, 1,500 warmup iterations, 1,500 sampling iterations. |
| Adapt delta | – | 0.999. |
| Maximum treedepth | – | 30. |
| Random seed | – | 20260519. |
| Exported posterior quantities | – | Incubation-period PMF, observed-pair generation-interval PMF, pair-level incubation periods, observed-pair generation intervals, Δ values, and serial intervals. |

### Supplementary Method 1. Latent infection/onset-time model

This section describes the within-host-informed latent infection/onset-time model used to estimate infection times, symptom-onset times, incubation periods, generation intervals, serial intervals, and transmission timing relative to source symptom onset in the Epuyén Andes-virus transmission-pair data. The model combines two components. First, it retains the continuous latent infection/onset-time structure of the Epiforecasts Epuyén Andes-virus line-list framework [12], which estimates infection and symptom-onset times within interval-censored exposure and onset windows, derives epidemiological timing quantities from those latent times, and models observed offspring counts using a time-varying reproduction number with negative-binomial dispersion. Second, it replaces the original heuristic delay distributions in that framework with a reduced observed-pair implementation of the within-host-informed event-time framework developed in [15].

The companion event-time framework treats epidemiological delays as linked outcomes of a shared within-host process, in which pathogen-load-driven event hazards can generate coupled latent-period, incubation-period, generation-interval, and serial-interval distributions [15]. In the present ANDV analysis, the available data consist of interval-censored exposure and onset windows plus reconstructed transmission links, but not individual viral-load measurements, detailed contact histories, or non-transmission risk sets. We therefore used a reduced implementation appropriate for linked transmission-pair timing data: symptom onset was modelled using a first-event hazard driven by a shared relative pathogen-burden trajectory, while linked-pair generation timing was regularized using a normalized observed-pair transmission-intensity model. This implementation was designed to estimate epidemiological timing distributions, not to infer individual viral loads, absolute transmission hazards, contact-tracing completeness, quarantine effectiveness, or intervention effects.

The present model should therefore be interpreted as a within-host-informed extension of the previous Epuyén latent-time line-list analysis [12], using the event-time logic developed in [15]. The latent-time structure, source–recipient transmission-pair structure, derived timing relationships, and offspring-count component follow the previous Epuyén framework, whereas the incubation-period and observed-pair generation-interval components are regularized by the shared within-host-informed timing model. We discuss the details of our implementation of [15]

#### Event-time structure

For case $i$, let $I_i$ denote infection time and $S_i$ denote symptom-onset time. The incubation period is defined as

$$IP_i = S_i - I_i.$$

For a linked transmission pair $i \to j$, where case $i$ infected case $j$, the generation interval is

$$GI_{ij} = I_j - I_i,$$

and the serial interval is

$$SI_{ij} = S_j - S_i.$$

Transmission timing relative to source symptom onset is

$$\Delta_{ij} = I_j - S_i.$$

Negative values of $\Delta_{ij}$ indicate transmission before inferred symptom onset of the source case, whereas positive values indicate transmission after source symptom onset.

These quantities satisfy

$$SI_{ij} = GI_{ij} + IP_j - IP_i.$$

Thus, the serial interval was treated as a derived quantity arising from latent infection and symptom-onset times, rather than as a primitive delay distribution. This construction preserves the deterministic relationships among incubation periods, generation intervals, serial intervals, and transmission timing relative to source symptom onset.

#### Latent infection and onset times

The model followed a latent-time approach in which infection and symptom-onset times were estimated for each case within case-specific time windows. Interval-censored symptom-onset information defined an onset interval

$$S_i \in [S_i^L, S_i^U],$$

and exposure information defined an infection-time interval

$$I_i \in [E_i^L, E_i^U].$$

For cases without complete exposure-window information, broad fallback windows were used relative to onset time or source symptom onset time. These fallback windows allowed model fitting but represented weaker timing information than observed exposure intervals.

The Stan model [16] estimated the latent infection and onset times through constrained latent variables. Posterior draws of $I_i$ and $S_i$ were then transformed into case-level incubation periods and pair-level generation intervals, serial intervals, and $\Delta$ values.

#### Within-host-informed timing model

The delay distributions were regularized using a shared within-host-informed event-time model [15]. Let $V(t)$ denote relative pathogen burden at time $t$ since infection. We used a rise-and-fall trajectory,

$$V(t) = A\exp(rt)\left\{1 + \frac{r}{d}\exp[\kappa(t-m)]\right\}^{-(r+d)/\kappa},$$

where $r > 0$ controls early growth, $d > 0$ controls late decline, $m$ controls approximate peak timing, $\kappa > 0$ controls the sharpness of the transition from growth to decline, and $A > 0$ is an arbitrary scale. Because the absolute pathogen-load scale was not identified from line-list data, $V(t)$ was used only as a relative trajectory shape.

Symptom onset was modelled as a first-event process with hazard proportional to this relative trajectory,

$$h_{\text{sym}}(t) = \lambda_{\text{sym}} V(t),$$

where $\lambda_{\text{sym}} > 0$ controls the timescale of symptom-onset activation. This implies an incubation-period density,

$$f_{IP}(t) = h_{\text{sym}}(t) \exp\left\{ - \int_0^t h_{\text{sym}}(u)\, du \right\},$$

which was normalized over the finite integration grid used in model fitting.

Linked-pair generation timing was regularized using a normalized observed-pair transmission-intensity model. The unnormalized observed-pair transmission intensity was proportional to the same relative trajectory,

$$w_{\text{trans}}(t) \propto V(t),$$

optionally multiplied by a soft transmission-timing gate. The observed-pair generation-interval density was obtained by normalizing this intensity over finite support,

$$f_{GI}^{\text{obs}}(t) = \frac{w_{\text{trans}}(t)}{\int_0^{t_{\max}} w_{\text{trans}}(u)\, du}.$$

This density describes the timing of realised linked transmission pairs conditional on transmission being observed. It should not be interpreted as a first-event transmission hazard, an absolute infectiousness profile, or a household risk-set model.

#### Transmission-pair likelihood

For each case $i$, the inferred incubation period,

$$IP_i = S_i - I_i,$$

contributed to the likelihood through the symptom-onset first-event density $f_{IP}(t)$. For each linked transmission pair $i \to j$, the inferred infection times imply

$$GI_{ij} = I_j - I_i.$$

The generation interval contributed to the likelihood through the normalized observed-pair generation-interval density $f_{GI}^{\text{obs}}(t)$. Transmission pairs with non-positive generation intervals were assigned zero probability.

The model also included the observed offspring count for each case. If $Z_i^{\text{obs}}$ is the number of observed secondary cases attributed to source $i$, then

$$Z_i^{\text{obs}} \sim \text{NegBin}_2(R_i, k_{\text{NB}}),$$

where $R_i$ is a time-varying reproduction number evaluated at the source case's onset time and $k_{\text{NB}}$ is the negative-binomial offspring-dispersion parameter. This offspring-count component was retained from the previous Epuyén latent-time line-list framework [12].

For post-control source cases, the primary model included a selection correction for observed-pair transmission timing under the normalized transmission-intensity model. Transmission events occurring after a specified post-onset isolation delay were downweighted by a residual contact probability, and the transmission-timing likelihood was renormalized. This correction accounted for the fact that observed post-control transmission pairs may reflect transmission under isolation and quarantine conditions rather than the unmitigated observed-pair timing distribution.

##### Derived posterior quantities and export to branching-process simulations

The Stan generated-quantities block exported posterior draws of the latent-time-derived quantities used in the containment simulations. For each posterior draw, case-level incubation periods and linked-pair generation intervals, serial intervals, transmission timings relative to source symptom onset were computed from the same sampled latent infection and onset times:

$$IP_i = S_i - I_i, \qquad GI_{ij} = I_j - I_i, \qquad \Delta_{ij} = I_j - S_i, \qquad SI_{ij} = S_j - S_i.$$

Thus, the exported pair-level incubation periods, generation intervals, transmission-timing values, and serial intervals preserve their posterior dependence rather than representing independent marginal draws timing quantities preserved the posterior dependence induced by the shared latent infection and symptom-onset times.

For the primary containment branching-process simulations, these posterior outputs were used as a fitted timing kernel for synthetic transmission trees. Each simulated transmission event sampled a row-level joint posterior timing draw. The sampled transmission timing relative to source symptom onset was combined with the simulated source incubation period to determine the generation interval, and the recipient incubation period was taken from the same posterior row. This avoided independently sampling incubation periods and generation intervals from marginal distributions, while allowing the simulations to generate new transmission trees that did not need to correspond one-to-one to the observed Epuyén transmission pairs.

The time-delay model did not directly estimate an absolute transmission-hazard scale, a contact process, or a separate infectiousness-onset time. Linked-pair generation timing was inferred from the normalized observed-pair transmission-intensity shape, whereas observed offspring counts were modelled separately through the negative-binomial $R_i$ component.

#### Supplementary Method 2. Stochastic branching-process contact-tracing model

##### Overview of the branching process

We used a stochastic individual-level branching-process model to evaluate quarantine capture, contact-tracing completeness, and post-response outbreak risk for person-to-person ANDV transmission under the Epuyén-calibrated higher-transmission and pooled lower-transmission scenarios.

The primary cruise-response analysis used a fixed public-health response day and a two-pass simulation procedure. A separate generation-start sensitivity analysis varied the generation at which tracing and quarantine began under the higher-transmission parameterization. The model was used for scenario analysis rather than forecasting, and the $R = 2.12$ parameterization should not be interpreted as the typical reproductive number of endemic ANDV transmission.

Each simulation began with $n_0 = 1$ introduced infection and generated a stochastic infection tree. The introduced infection was assigned transmission generation $g = 0$, and each retained secondary infection was assigned generation $g_j = g_i + 1$. Full assumptions and parameter values for the branching-process model are listed in Supplementary Table 4.

Disease-timing quantities used in the branching-process simulations were taken from the posterior outputs of the latent infection/onset-time model described in Supplementary Method 1. Each simulated infection was assigned an incubation period, and each potential transmission was assigned a generation interval, by sampling from the posterior-derived empirical timing outputs exported by that model. These sampled quantities determined symptom-onset times and proposed recipient infection times in the branching process. The branching-process model therefore did not re-estimate incubation-period, generation-interval, serial-interval, or transmission-timing distributions; these entered only as external posterior-derived inputs.

For each infected individual, the number of potential secondary infections was sampled from the scenario-specific overdispersed offspring distribution specified in Supplementary Table 4. In the primary containment analysis, the higher-transmission scenario used the Epuyén-calibrated parameterization $R = 2.12$ and $k = 0.316$, whereas the lower-transmission scenario used the pooled multi-study parameterization $R = 0.242$ and $k = 0.400$. This generated the uncontrolled potential transmission tree before isolation, quarantine, and tracing rules were applied. The negative-binomial offspring distribution followed the parameterization

$$E[Z_i^{\mathrm{pot}}] = R, \qquad \mathrm{Var}(Z_i^{\mathrm{pot}}) = R + \frac{R^2}{k},$$

consistent with standard branching-process representations of individual-level transmission heterogeneity and superspreading [11, 13]. Each potential secondary infection $i \to j$ was assigned a proposed infection time by adding a sampled posterior-derived generation interval to the source infection time. If the potential transmission was not prevented by the intervention rules during the replay step, it was retained and recipient $j$ was added to the realised simulated infection tree with generation $g_j = g_i + 1$. Potential transmissions were discarded if they occurred while the relevant individual was effectively quarantined or isolated under the scenario assumptions.

#### Disease-timing inputs

Disease-timing inputs were read from the posterior-derived incubation-period and generation-interval outputs of the latent infection/onset-time model described in Supplementary Method 1. For each simulated case, a sampled incubation period determined symptom-onset time relative to infection.

For each potential transmission edge, one row was sampled from the joint posterior timing draws. The generation interval and recipient incubation period were taken from that same posterior row, preserving their row-level dependence. The proposed recipient infection time was obtained by adding the sampled generation interval to the source infection time, and the recipient symptom-onset time was obtained by adding the sampled recipient incubation period to the recipient infection time.

#### Transmission generation, current-generation conditioning, and delayed-start scenarios

The branching process explicitly tracked transmission generation. The introduced case was assigned generation $g = 0$, and each retained secondary infection from source $i$ to recipient $j$ was assigned generation

$$g_j = g_i + 1.$$

For the primary cruise-response analysis, the public-health response was anchored at a fixed time,

$$t_{\text{response}} = 35$$

days after the simulation origin. Simulations were conditioned on the uncontrolled potential transmission tree reaching generation 2 by this response day. Future outcomes were then summarized after the response day and after the current generation, denoted

$$g_{\text{current}} = 2.$$

Cases with $g_i > g_{\text{current}}$ were counted as additional infections after the current generation.

In a separate generation-start sensitivity analysis, we varied the generation at which tracing and quarantine began:

$$g_{\text{start}} \in \{0, 1, 2, 3, 4\}.$$

This analysis was separate from the primary cruise day-35 response analysis and was used to assess how delayed initiation of tracing and quarantine affected outbreak size, quarantine burden, and response duration.

#### Case detection and isolation

Each infected individual was detected with probability

$$D_i \sim \text{Bernoulli}(p_{\text{det}}),$$

with baseline value

$$p_{\text{det}} = 0.95.$$

If detected, detection occurred after symptom onset:

$$T_i^{\text{det}} = S_i + \delta_i,$$

where

$$\delta_i \sim \text{Gamma}\left(\alpha_{\text{det}} = 4,\ \beta_{\text{det}} = 2\right),$$

using a shape–rate parameterization. This distribution has mean 2.0 days and SD 1.0 day. Detected cases were isolated after an additional delay,

$$T_i^{\text{iso}} = T_i^{\text{det}} + \epsilon_i,$$

where

$$\epsilon_i \sim \text{Gamma}\left(\alpha_{\text{iso}} = 1,\ \beta_{\text{iso}} = 2\right),$$

using a shape–rate parameterization. This distribution has mean 0.5 days and SD 0.5 days.

In the generation-start sensitivity analysis, detection and isolation could reduce onward transmission only after the scenario-specific response time, defined by the first infection in generation $g_{\text{start}}$. If a potential transmission from source $i$ to recipient $j$ was scheduled at time $I_j$, the residual probability of the scheduled transmission surviving source isolation was

$$r_i^{\text{iso}}(I_j) = \begin{cases} 1, & I_j < t_{\text{response},r}(g_{\text{start}}), \\ 1, & I_j \geq t_{\text{response},r}(g_{\text{start}}) \text{ and } I_j < T_i^{\text{iso}}, \\ 1 - e_{\text{iso}}, & I_j \geq t_{\text{response},r}(g_{\text{start}}) \text{ and } I_j \geq T_i^{\text{iso}}. \end{cases}$$

The baseline isolation effectiveness was

$$e_{\text{iso}} = 0.99.$$

#### Contact model for tracing

In the primary fixed-response cruise analysis, for each detected source case we constructed a traceable contact pool over the overlap between the backward tracing window and the period from infection to detection. The start of this traceable period was

$$T_i^{\text{trace,start}} = \max(S_i - b, I_i),$$

where $b$ is the backward tracing window. The end of the traceable period was

$$T_i^{\text{trace,end}} = T_i^{\text{det}}.$$

The traceable contact duration was therefore

$$d_i = \max\left(0, T_i^{\text{trace,end}} - T_i^{\text{trace,start}}\right).$$

Total contacts over this period were drawn from a negative-binomial contact distribution informed by social-contact survey data [21, 22]:

$$C_i \sim \mathrm{NegBin}(\mu_C d_i, k_C d_i),$$

where $\mu_C$ is the mean number of contacts per day and $k_C$ is the contact-count dispersion parameter. In the baseline model,

$$\mu_C = 13.495956, \qquad k_C = 2.114096.$$

The number of infected contacts in this pool was the number of realised infected offspring of source $i$ whose infection times fell within the traceable contact window:

$$Z_i^{\mathrm{trace}} = \sum_j \mathbf{1}\left(I_j \in [T_i^{\mathrm{trace,start}}, T_i^{\mathrm{trace,end}}]\right),$$

where the sum is over realised infected recipients of source $i$. The total contact count was constrained to be at least $Z_i^{\mathrm{trace}}$. The number of uninfected contacts was then

$$U_i = C_i - Z_i^{\mathrm{trace}}.$$

#### Tracing completeness and selection of quarantined contacts

The main scenario parameter was the system tracing-completeness multiplier,

$$q \in [0, 1].$$

The default values were

$$q \in \{1.00, 0.95, 0.90, 0.75, 0.50\}.$$

In the primary fixed-response tracing model, each realised infected contact $j$ whose infection time fell within the traceable period was successfully identified independently with probability $q$:

$$B_{ij}^{\mathrm{trace}} \sim \mathrm{Bernoulli}(q).$$

If $Z_i^{\mathrm{trace}}$ denotes the number of realised infected contacts whose infection times fell within the traceable period, the reconstructed number of uninfected contacts was

$$U_i = C_i - Z_i^{\mathrm{trace}}.$$

The number of traced uninfected contacts was then sampled as

$$N_i^{\mathrm{trace,uninf}} \sim \mathrm{Binomial}(U_i, q).$$

Thus, the same tracing-completeness parameter $q$ governed independent identification of realised infected contacts and tracing of contacts who were retrospectively uninfected.

#### Quarantine

The model included two quarantine mechanisms: quarantine of the known exposed cruise cohort at the fixed public-health response time, and quarantine of traced contacts identified through subsequent contact tracing.

In the primary cruise-response analysis, the public-health response was anchored at

$$t_{\text{response}} = 35$$

days after the simulation origin. All infected individuals with infection time at or before $t_{\text{response}}$ were treated as infected members of the known exposed cohort at response. Each infected cohort member was captured into quarantine independently with probability $q$. Captured infected cohort members entered quarantine for 42 days.

The model also tracked uninfected members of the exposed cruise cohort. If the known exposed cohort size was $N_{\text{cohort}}$, and $N_{\text{inf,response}}$ infected cohort members were present at response, then the number of uninfected exposed individuals was

$$N_{\text{uninf,response}} = \max\left(0, N_{\text{cohort}} - N_{\text{inf,response}}\right).$$

These uninfected exposed individuals contributed to quarantine burden but not to transmission. Each uninfected exposed individual was quarantined independently with probability $q$.

For each successfully traced infected contact $j$ identified from source $i$, the notification delay was sampled as

$$\omega_{ij} \sim \text{Gamma}\left(\alpha_{\text{notify}} = 4,\ \beta_{\text{notify}} = 4\right),$$

using a shape–rate parameterization. This distribution has mean 1.0 day and SD 0.5 days. Quarantine began at

$$T_j^Q = T_i^{\text{det}} + \omega_{ij}.$$

Quarantine lasted 42 days:

$$T_j^{Q,\text{end}} = T_j^Q + 42.$$

For a gamma distribution parameterized by shape $\alpha$ and rate $\beta$, the mean is $\alpha/\beta$ and the standard deviation is $\sqrt{\alpha}/\beta$.

In the baseline analysis, quarantine adherence was 1.0 and quarantine effectiveness was

$$e_Q = 1.0.$$

The residual transmission probability during quarantine was

$$r_j^Q(t) = \begin{cases} 1, & t < T_j^Q \text{ or } t > T_j^{Q,\text{end}}, \\ 1 - e_Q, & T_j^Q \le t \le T_j^{Q,\text{end}}. \end{cases}$$

The model recorded whether traced infected contacts entered quarantine before symptom onset,

$$T_j^Q \le S_j.$$

It did not classify contacts according to whether they were quarantined before infectiousness onset, because infectiousness onset was not explicitly represented in the branching-process model.

##### Intervention replay and retained transmissions

In the primary cruise-response analysis, simulations were run in two passes. First, an uncontrolled potential transmission tree was generated. Second, potential transmissions were replayed chronologically after imposing isolation, quarantine, and contact-tracing rules from the fixed response day. Transmissions that survived these intervention rules were retained. These retained transmissions defined the realised simulated outbreak and were counted in the main-text outcomes as continued transmission, additional infections after generation 2, and total infections.

Potential transmissions that did not survive because the source or recipient was isolated or quarantined were counted as prevented transmissions. For each potential secondary infection $i \to j$, the model evaluated whether the scheduled infection at time $I_j$ survived source isolation and quarantine under the scenario assumptions. The combined residual transmission probability was

$$r_i(I_j) = r_i^{\text{iso}}(I_j)\, r_i^Q(I_j).$$

The potential transmission was retained with probability $r_i(I_j)$. If retained, the recipient infection was added to the infection tree; if not retained, the transmission was counted as prevented by isolation or quarantine.

The realised offspring count for individual $i$ was

$$Z_i^{\text{real}} = \sum_j \mathbf{1}\left(\text{potential transmission } i \to j \text{ was retained}\right).$$

##### Current-generation and delayed-start outcomes

To align simulations with the current outbreak context, primary cruise-response analyses were conditioned on the uncontrolled potential transmission tree reaching generation 2 by the fixed response day. Let $g_{\text{current}} = 2$, and let

$$\mathcal{R}_q = \{r : \exists i \text{ such that } g_{ir} = g_{\text{current}} \text{ and } I_{ir} \le t_{\text{response}}\}$$

denote the set of stochastic replicates under tracing completeness $q$ that reached generation 2 by the response day. Let

$$B_q^{(2)} = |\mathcal{R}_q|.$$

Additional post-response infections were defined as

$$A_{\text{post},r}(q) = \sum_i \mathbf{1}\left\{I_{ir} > t_{\text{response}}\right\},$$

where $I_{ir}$ is the realised infection time of case $i$ in replicate $r$.

For Figure 2A, continued transmission was defined as at least one post-response transmission occurring outside quarantine and was estimated across all simulations in $\mathcal{R}_q$.

For Figure 2B, the number of additional post-response infections was summarized only among simulations satisfying

$$A_{\text{post},r}(q) > 0.$$

For the separate delayed-start and current-generation sensitivity analyses, additional infections after the current generation were defined as

$$A_{\text{gen},r}(q) = \sum_i \mathbf{1}\left\{g_{ir} > g_{\text{current}}\right\}.$$

This generation-based outcome was not the conditioning variable used for Figure 2B.

For Figures 2C and 2D, downstream quarantine burden and additional response duration were summarized across all simulations in $\mathcal{R}_q$, including simulations with no additional post-response infection.

For the primary Figure 2 analysis, total response duration was defined as the interval from the fixed day-35 response until completion of all subsequent simulated transmission, detection, tracing, and quarantine activity. Additional response duration beyond the initial quarantine period was defined as

$$D_{\text{additional},r} = \max\left\{0,\ D_{\text{total},r} - 42 \text{ days}\right\}.$$

Additional response duration was converted to months using 30.4375 days per month and was summarized across all simulations satisfying the generation-2 conditioning.

For the delayed-start sensitivity analysis, simulations were repeated across

$$g_{\text{start}} \in \{0, 1, 2, 3, 4\}.$$

For each delayed-start scenario, the response point was defined as the earliest infection time among cases in the specified start generation:

$$t_{\text{response},r}(g_{\text{start}}) = \min_i \left\{I_{ir} : g_{ir} = g_{\text{start}}\right\}.$$

At this scenario-specific response point, infected individuals already present in the simulated outbreak, including infections in earlier generations, were eligible for capture and quarantine with probability $q$. Subsequent detected post-response cases could also generate contact tracing and quarantine according to the same tracing-completeness parameter.

Additional quarantine burden in the delayed-start analysis was summarized from the scenario-specific response point onward. This burden included infected individuals captured at the response point or later traced and quarantined, plus retrospectively uninfected contacts quarantined through post-response tracing activity:

$$A_{Q,\text{total},r} = A_{Q,\text{inf},r} + A_{Q,\text{uninf},r}.$$

Here, $A_{Q,\text{inf},r}$ counts infected individuals quarantined through the response or subsequent tracing, and $A_{Q,\text{uninf},r}$ counts retrospectively uninfected contacts quarantined through post-response tracing. The inclusion of earlier-generation infected individuals at the response point reflects the assumed start of tracing and quarantine in that scenario; it does not mean that all detected generation-1 cases were retrospectively traced only because generation 2 had begun.

As with the full-outbreak quarantine summaries, $A_{Q,\text{uninf},r}$ represents precautionary quarantine burden assessed retrospectively. It does not imply that quarantine was unnecessary at the time of decision-making, because infection status is unknown prospectively.

For the delayed-start sensitivity analysis, response duration was defined as the time from the scenario-specific response point until the end of all subsequent simulated transmission, detection, tracing, and quarantine activity.

For each $g_{\text{start}}$, outcomes were summarized using the probability of any additional cases after generation 2, the number of additional infections after generation 2, total infections, additional quarantine burden after the scenario-specific response point, and response duration after the scenario-specific response point. Summary statistics included the 95th percentile of total infections, the 95th percentile of additional quarantine burden, and the 95th percentile of response duration after the scenario-specific response point.

For each replicate and scenario, recorded run-level outcomes included the tracing-start and current generations, whether each was reached, the scenario-specific response time, total potential and retained infections, the number and occurrence of additional cases after generation 2, post-response infections and transmissions, additional quarantine burden, transmissions prevented by quarantine or isolation, and response duration after the scenario-specific response point.

#### Real-time inference of transmission from accumulating detected cases

To evaluate how quickly the transmission scenario could be inferred from information available early in an outbreak, we performed a separate real-time inference analysis using the same two transmission parameterizations as the main containment analyses: the Epuyén-calibrated higher-transmission scenario ($R = 2.12$, $k = 0.316$) and the pooled lower-transmission scenario ($R = 0.242$, $k = 0.400$). For each scenario, we simulated 10,000 uncontrolled potential transmission trees using the same posterior-derived disease-timing distributions, case-detection probability, and reporting-delay assumptions used in the main branching-process model.

We evaluated inference when the simulated outbreak accumulated

$$n \in \{1, 2, 3, 5, 10, 15, 20\}$$

detected cases. At each threshold, the simulated outbreak was observed immediately before an intervention would begin, and only information that would have been available by that time was used for inference. Analyses at a given threshold were therefore conditional on the simulated outbreak having reached that number of detected cases.

For each detected source case $i$, let $y_i(t)$ denote the number of its direct secondary infections that had themselves been detected by observation time $t$. Because offspring infected shortly before the observation time may not yet have developed symptoms or been detected, the observed offspring count is right truncated. We accounted for this using

$$a_i(t) = p_{\text{det}} P\left(T_{\text{det},j} \leq t \mid S_i\right),$$

where $p_{\text{det}} = 0.95$, $S_i$ is the symptom-onset time of source $i$, and the distribution of recipient detection time relative to source symptom onset was obtained from the posterior-derived serial-interval distribution combined with the assumed symptom-onset-to-detection delay.

Under binomial thinning of the negative-binomial offspring distribution, the observed number of detected offspring was modelled as

$$y_i(t) \sim \text{NegBin}_2\left(R a_i(t),\ k\right).$$

Thus, sources with little follow-up time contributed less information than sources whose potential secondary cases had sufficient time to become detectable.

At each simulated outbreak and detected-case threshold, $R$ and $k$ were estimated jointly using weakly regularising priors on the log scale,

$$\log R \sim \text{Normal}\left(\log 1,\ 1.5\right),$$

and

$$\log k \sim \text{Normal}\left(\log 0.3,\ 1.25\right).$$

The posterior mode was obtained numerically on the $(\log R, \log k)$ scale. Approximate within-outbreak posterior uncertainty was then obtained from the local curvature of the log posterior using a Laplace approximation. For each outbreak-threshold combination, we calculated the posterior median and 95% credible interval for $R$, together with the posterior probability

$$P(R > 1).$$

Across simulated outbreaks reaching each threshold, the central estimate shown in Supplementary Figure S1 is the median of the within-outbreak posterior medians. The uncertainty band represents the median lower and upper limits of the within-outbreak 95% credible intervals and therefore summarizes typical inferential uncertainty for an individual outbreak; it is not the between-simulation spread of point estimates. Thresholds reached by fewer than 20 of the 10,000 simulations were omitted from the plotted summaries.

Because results at each threshold are conditional on attaining that case count, lower-transmission outbreaks reaching several detected cases form an increasingly selected subset with unusually high realised transmission. Estimates at later thresholds under the lower-transmission parameterization should therefore not be interpreted as estimates for a typical outbreak generated with $R = 0.242$, but rather as inference from the rare lower-transmission outbreaks that happened to grow sufficiently large to be observed at those thresholds.

#### Supplementary Method 3. Compatibility of the 2026 cruise-associated outbreak with transmission scenarios

**Observed cruise-associated outbreak data.** The observed epidemic trajectory was constructed from symptom-onset dates for the 13 reported cases associated with the 2026 multinational cruise-ship outbreak. The symptom-onset dates used in this analysis were 3 April, 21 April, 22 April, 23 April, 28 April (two cases), 30 April, 1 May, 8 May, 11 May, 14 May, 20 May, and 30 May 2026. For comparison with simulated outbreaks, calendar dates were converted to elapsed time relative to the earliest observed symptom onset, such that 3 April 2026 corresponded to day 0. The resulting onset times were aggregated into 7-day intervals to form the observed weekly epidemic curve.

**Transmission scenarios and outbreak simulation.** We assessed the compatibility of the observed epidemic curve with the two transmission scenarios used in the main analysis. The higher-transmission scenario used a negative-binomial offspring distribution with mean reproduction number $R = 2.12$ and dispersion parameter $k = 0.316$, calibrated to the pre-intervention Epuyén outbreak. The lower-transmission scenario used the pooled multi-study estimates $R = 0.242$ and $k = 0.400$. For each scenario, we generated 10,000 independent stochastic outbreaks beginning with a single introduced infection.

For each infected individual, the number of secondary infections was drawn from the corresponding negative-binomial offspring distribution. Generation intervals were sampled from the posterior-derived generation-interval distribution estimated by the latent infection/onset-time model, and recipient incubation periods were sampled from the corresponding posterior-derived incubation-period distribution. Proposed recipient infection times were obtained by adding the sampled generation interval to the source infection time, and symptom-onset times were obtained by adding the sampled incubation period to each recipient infection time. For this separate epidemic-curve compatibility analysis, simulations were terminated at 5,000 cases or 150 days after introduction; these computational safeguards are distinct from those used for the containment analyses summarized in Table 3.

Simulated symptom-onset trajectories were aligned to the first symptom onset within each outbreak and aggregated into the same 7-day intervals used for the observed data.

**Epidemic-curve compatibility statistic.** For each transmission scenario, let $\mathbf{X}_s$ denote the vector of weekly case counts in simulated outbreak $s$, and let $\mathbf{x}_{\text{obs}}$ denote the corresponding observed weekly incidence vector. Weeks with zero variance across all simulated outbreaks under a given scenario were excluded because they contained no information for distinguishing the observed trajectory from simulations. We calculated the simulation mean vector

$$\boldsymbol{\mu} = \frac{1}{N}\sum_{s=1}^{N}\mathbf{X}_s$$

and covariance matrix

$$\boldsymbol{\Sigma} = \text{Cov}(\mathbf{X}_1, \ldots, \mathbf{X}_N),$$

where $N = 10{,}000$. To ensure numerical stability when inverting the covariance matrix, a small ridge term was added to its diagonal. Specifically,

$$\boldsymbol{\Sigma}^* = \boldsymbol{\Sigma} + \lambda\mathbf{I},$$

where

$$\lambda = \max\left\{10^{-6}, 10^{-4}\,\text{mean}[\text{diag}(\boldsymbol{\Sigma})]\right\}.$$

Compatibility of the observed temporal trajectory with each transmission scenario was summarized using its squared Mahalanobis distance from the simulated mean,

$$D_{\text{obs}} = (\mathbf{x}_{\text{obs}} - \boldsymbol{\mu})^T(\boldsymbol{\Sigma}^*)^{-1}(\mathbf{x}_{\text{obs}} - \boldsymbol{\mu}).$$

For comparison, the same statistic was calculated for each simulated epidemic curve,

$$D_s = (\mathbf{X}_s - \boldsymbol{\mu})^T(\boldsymbol{\Sigma}^*)^{-1}(\mathbf{X}_s - \boldsymbol{\mu}).$$

We then calculated an empirical compatibility probability as

$$p_{\text{compat}} = \frac{1}{N}\sum_{s=1}^{N}\mathbb{I}(D_s \geq D_{\text{obs}}).$$

Thus, small values of $p_{\text{compat}}$ indicate that the observed weekly epidemic trajectory lies unusually far from trajectories generated under that transmission scenario. We treated $p_{\text{compat}} < 0.05$ as evidence of poor compatibility, while larger values indicated that the observed trajectory was not unusual relative to simulations from that scenario. This quantity is a simulation-based model-compatibility measure rather than a frequentist test of one transmission scenario against the other.

**Compatibility with observed outbreak size.** Because temporal similarity alone does not capture the probability that a transmission scenario generates an outbreak as large as that observed, we additionally calculated

$$P(N_{\text{cases}} \geq 13)$$

as the proportion of simulated outbreaks containing at least 13 infections. This quantity describes how frequently an introduction under each transmission parameterization generated an outbreak at least as large as the 13-case cruise-associated cluster. The trajectory-based and outbreak-size summaries were interpreted jointly rather than as formal posterior probabilities for the two competing transmission scenarios.

### Supplementary Results

#### Supplementary timing estimates

To aid interpretation of the timing estimates reported in the main text, we plotted the model-implied timing distributions from the latent infection/onset-time model and summarized fitted delay distributions using the reporting framework of Charniga et al [8]. These summaries include central tendency, posterior uncertainty, fitted distributional spread, and upper-tail quantiles rather than only point estimates of mean timing values.

Figure 1 shows the fitted timing distributions. The solid line shows the posterior median density, and the shaded ribbon shows posterior uncertainty in the estimated density curve. Supplementary Table 6 reports posterior uncertainty in the mean, median, and SD of each delay distribution, along with fitted IQRs, central 95% distributional intervals, and upper-tail quantiles.

**Table 6: Estimated delay distributions from the within-host-informed latent infection/onset-time model.** Mean, median, and SD estimates are posterior medians with 95% credible intervals. IQRs, central 95% intervals, and upper-tail quantiles summarize fitted individual-level variation across infections or linked transmission pairs. Transmission timing is defined relative to source symptom onset, with negative values indicating transmission before source symptom onset.

| Quantity | Mean | Median | SD | IQR | Central 95% interval | 95th / 99th percentile |
|---|---|---|---|---|---|---|
| Incubation period | 19.36 (17.56–20.99) | 18.50 (16.50–19.50) | 6.95 (5.92–8.21) | 14.50–23.50 | 7.50–35.50 | 31.50 (29.50–35.50) / 39.50 (35.50–44.50) |
| Latent period | 13.52 (11.09–15.91) | 13.50 (10.50–15.50) | 4.44 (3.26–6.01) | 10.50–16.50 | 5.50–22.50 | 20.50 / 25.50 |
| Generation interval | 23.16 (21.49–25.05) | 22.50 (20.50–23.50) | 6.83 (5.75–8.17) | 18.50–26.50 | 12.50–38.50 | 35.50 / 43.50 |
| Serial interval | 22.64 (22.42–22.83) | 21.79 (21.28–22.18) | 7.06 (6.92–7.15) | 18.93–24.71 | 12.20–36.94 | 36.05 / 38.96 |

*Continued on next page*

| Quantity | Mean | Median | SD | IQR | Central 95% interval | 95th / 99th percentile |
|---|---|---|---|---|---|---|
| Transmission timing relative to source symptom onset | 0.12 (−0.05–0.38) | 0.07 (−0.20–0.30) | 0.80 (0.67–1.01) | −0.25–0.39 | −0.85–2.02 | 1.23 / 2.78 |

##### Delay-distribution reporting checklist

To make the delay estimates transparent and reproducible, we assessed the within-host-informed latent infection/onset-time model against the delay-distribution reporting recommendations of Charniga et al [8]. Supplementary Table 7 summarizes where each reporting item is addressed in the manuscript or Supplementary Information.

**Table 7: Delay-distribution reporting checklist for the within-host-informed latent infection/onset-time model.** Checklist items were adapted from the reporting recommendations of Charniga et al [8].

| Checklist item | Addressed | Location / notes |
|---|---|---|
| Delay distribution clearly named and defined | Yes | Supplementary Results define incubation period, latent period, generation interval, serial interval, and transmission timing relative to source symptom onset. |
| Biological or epidemiological interpretation of each delay given | Yes | Incubation period is time from infection to symptom onset; latent period is the model-implied time from infection to infectiousness activation; generation interval is source infection to recipient infection; serial interval is source symptom onset to recipient symptom onset; $\Delta_{ij}$ is recipient infection time relative to source symptom onset. |
| Data source and outbreak context described | Yes | Main Methods and Supplementary Method 1 describe the 2018–2019 Epuyén ANDV transmission-pair line list. |
| Case inclusion criteria and sample size reported | Yes | Main Methods and Supplementary Method 1 report the cases and linked transmission pairs used in the latent-time model. |
| Exposure and event-time censoring described | Yes | Supplementary Method 1 describes interval-censored exposure and symptom-onset windows, with latent infection and onset times estimated within those windows. |
| Treatment of missing or uncertain exposure windows described | Yes | Supplementary Method 1 and Supplementary Table 5 describe broad fallback infection-time windows used when complete exposure-window information was unavailable. |
| Transmission-pair structure described | Yes | Main Methods and Supplementary Method 1 describe use of reconstructed source–recipient links from the Epuyén outbreak. |
| Multiple possible infectors or source uncertainty described | Yes | Supplementary Table 5 describes the source-link policy used for cases with multiple listed possible sources. |
| Model form and parametric assumptions stated | Yes | Supplementary Method 1 describes the within-host-informed event-time model, including the pathogen-burden trajectory, symptom-onset hazard, observed-pair generation-interval model, and offspring-count model. |
| Whether estimates are fitted distributions or empirical observed intervals clarified | Yes | Supplementary Results and Figure 1 state that plotted curves are model-implied fitted timing distributions across infections or linked transmission pairs. |

*Continued on next page*

| Checklist item | Addressed | Location / notes |
|---|---|---|
| Central tendency reported | Yes | Supplementary Table 6 reports posterior estimates of the mean and median for each delay distribution. |
| Distributional variability reported | Yes | Supplementary Table 6 reports SDs, IQRs, and central 95% fitted distributional intervals. |
| Upper-tail quantiles reported | Yes | Supplementary Table 6 reports 95th and 99th percentiles for each delay distribution. |
| Uncertainty intervals reported for estimated quantities | Yes | Supplementary Table 6 reports 95% credible intervals for the mean, median, and SD of each delay distribution. |
| Timing relative to symptom onset clearly signed | Yes | Transmission timing is defined as $\Delta_{ij} = I_j - S_i$, so negative values indicate transmission before source symptom onset. |
| Pre-symptomatic transmission probability reported | Yes | Main Results and Table 1 report the posterior estimate of the proportion of transmission occurring before source symptom onset. |
| Model diagnostics reported | Yes | Supplementary diagnostics report sampler diagnostics, including divergences, effective sample sizes, and $\hat{R}$ values. |
| Model fit or posterior predictive checks shown | Yes | Supplementary timing-distribution plots are shown in Figure 1. |
| Sensitivity analyses or alternative assumptions described | Yes | Supplementary Methods describe prior settings, model assumptions, control-selection correction, and scenario analyses used to assess sensitivity to response timing and tracing completeness. |
| Code and data availability provided | Yes | The Code availability statement provides the analysis repository and reproducibility materials. |
| Comparison with previous delay estimates included | Yes | The Supplementary comparison subsection and Supplementary Table 8 compare estimates with the previous latent-time Epuyén line-list analysis. |

##### Comparison with a previous latent-time re-analysis of the Epuyén line list

We compared epidemiological delay estimates from the within-host-informed latent infection/onset-time model with those from the previous latent-time re-analysis of the 2018–2019 Epuyén line list [12]. The previous analysis jointly estimated latent infection and symptom-onset times, incubation periods, transmission timing relative to source symptom onset, weekly time-varying reproduction numbers, and negative-binomial offspring dispersion from interval-censored transmission-pair data. In contrast, the present model used a within-host-informed event-time structure for incubation periods and linked-pair generation timing. The previous analysis used a lognormal incubation-period distribution and a normal distribution for transmission timing relative to source symptom onset, denoted $\Delta$. Generation and serial intervals were derived in post-processing from the fitted incubation-period and transmission-timing distributions.

The two analyses gave similar estimates for the timing of person-to-person transmission relative to symptom onset. The previous analysis estimated $\mu_\Delta = 0.17$ days after source symptom onset (95% CrI: $-0.16$ to 0.49), whereas the present model estimated mean $\Delta = 0.12$ days (95% CrI: $-0.05$ to 0.38) and median $\Delta = 0.07$ days (95% CrI: $-0.20$ to 0.30). Thus, both analyses placed transmission very close to source symptom onset. The present model estimated a somewhat wider distribution of $\Delta$, with SD 0.80 days (95% CrI: 0.67 to 1.01), compared with $\sigma_\Delta = 0.61$ days (95% CrI: 0.45 to 0.84) in the previous analysis.

The main difference between analyses was in the incubation-period distribution. The previous analysis estimated a mean incubation period of 22.49 days (95% CrI: 20.26 to 25.48), whereas the present model estimated a shorter mean incubation period of 19.36 days (95% CrI: 17.56 to 20.99) and median incubation period of 18.50 days (95% CrI: 16.50 to 19.50). The present model also estimated a shorter upper tail, with incubation-period 95th and 99th percentiles of 31.50 and 39.50 days, compared with 36.12 days (95% CrI: 31.31 to 44.10) and 44.95 days (95% CrI: 37.50 to 58.12), respectively, in the previous analysis.

Generation and serial-interval estimates were broadly similar across analyses. The previous analysis reported a combined generation/serial-interval mean of 22.68 days (95% CrI: 20.42 to 25.67) and SD of 7.38 days (95% CrI: 5.61 to 10.38). The present model estimated a mean generation interval of 23.16 days (95% CrI: 21.49 to 25.05) and mean serial interval of 22.64 days (95% CrI: 22.42 to 22.83). These comparisons suggest that the two approaches agree on the central timing of transmission, while differing mainly in the inferred incubation-period distribution and, consequently, the relative decomposition of infection-to-onset and infection-to-transmission timing.

**Table 8: Comparison of epidemiological delay estimates from the previous Epuyén latent-time line-list analysis and the present within-host-informed latent infection/onset-time model.** Values are posterior median estimates with 95% credible intervals unless otherwise stated. For the present analysis, distributional quantiles are posterior median fitted quantiles.

| **Quantity** | **Previous latent-time analysis** | **Present analysis** |
|---|---|---|
| Incubation-period mean | 22.49 (20.26–25.48) | 19.36 (17.56–20.99) |
| Incubation-period median | – | 18.50 (16.50–19.50) |
| Incubation-period SD | – | 6.95 (5.92–8.21) |
| Incubation-period central 95% interval | – | 7.50–35.50 |
| Incubation-period 95th percentile | 36.12 (31.31–44.10) | 31.50 |
| Incubation-period 99th percentile | 44.95 (37.50–58.12) | 39.50 |
| Transmission-timing mean, $\Delta$ | 0.17 (−0.16–0.49) | 0.12 (−0.05–0.38) |
| Transmission-timing median, $\Delta$ | – | 0.07 (−0.20–0.30) |
| Transmission-timing SD, $\Delta$ | 0.61 (0.45–0.84) | 0.80 (0.67–1.01) |
| Transmission-timing central 95% interval, $\Delta$ | Approx. −1.02–1.36[a] | −0.85–2.02 |
| Generation/serial-interval mean | 22.68 (20.42–25.67) | GI: 23.16 (21.49–25.05); SI: 22.64 (22.42–22.83) |
| Generation/serial-interval SD | 7.38 (5.61–10.38) | GI: 6.83 (5.75–8.17); SI: 7.06 (6.92–7.15) |
| Negative-binomial offspring dispersion, $k$ | 0.38 (0.16–1.07) | – |

[a]Derived from the posterior median estimates of $\mu_\Delta$ and $\sigma_\Delta$ under the previous analysis's normal transmission-timing model; this interval is shown only to aid comparison with the distributional interval reported for the present analysis.

#### Early inference of the reproduction number from detected cases

Inference about the reproduction number was highly uncertain during the first several detected cases (Supplementary Figure S1). Under both simulated transmission scenarios, the within-outbreak 95% credible intervals were broad at early case-count thresholds and commonly included the critical value

$R = 1$. Consequently, the first few detected cases provided limited evidence for distinguishing lower-transmission from higher-transmission.

Under the higher-transmission scenario, estimates became progressively more informative as additional detected cases accumulated, although substantial uncertainty remained during the early stages of the outbreak. In contrast, lower-transmission outbreaks that reached several detected cases produced estimated reproduction numbers substantially above the generating value of $R = 0.242$. This did not indicate bias toward higher-transmission in typical lower-transmission outbreaks. Rather, conditioning on attainment of multiple detected cases preferentially selected the small subset of highly overdispersed lower-transmission realizations that had experienced unusually high realised transmission.

This selection became increasingly important at later thresholds because lower-transmission outbreaks rarely accumulated large numbers of detected cases. Consequently, the lower-transmission trajectory could not be estimated reliably at the later case-count thresholds, whereas higher-transmission outbreaks continued to provide information as case numbers accumulated. Overall, these simulations show that an outbreak generated by a higher-transmission process may remain difficult to identify as higher-transmission from the first several detected cases alone. Conversely, observing several cases during a highly overdispersed lower-transmission outbreak can transiently resemble evidence for higher transmission. Early case counts therefore provide limited information for confidently excluding either transmission scenario.

##### Threshold probabilities for additional infections after generation 2

To complement the full distributional summaries of additional infections after the current generation-2 response point, we calculated exceedance probabilities for several operationally relevant thresholds. These are supplementary generation-based endpoints and are distinct from the primary Figure 2A outcome, which is defined as at least one post-response transmission occurring outside quarantine. Among simulations conditioned on the uncontrolled potential transmission tree reaching generation 2 by the fixed response day, we estimated the probability of no additional cases and the probabilities of at least 5, 10, 25, or 50 additional infections after generation 2 under each tracing-completeness scenario.

Under the Epuyén-calibrated higher-transmission scenario, these threshold summaries show that incomplete capture of the exposed cohort and incomplete downstream contact tracing increased not only the probability of any continued transmission, but also the probability of substantially larger downstream case burdens. Under complete tracing and quarantine capture, all conditioned simulations produced no additional cases after generation 2. At 95% tracing completeness, 80.4% of simulations produced no additional cases, whereas 15.9% produced at least 10 additional cases and 14.8% produced at least 50 additional cases. At 90% tracing completeness, the corresponding probabilities were 60.2%, 31.3%, and 28.6%. At 75% tracing completeness, they were 40.1%, 51.0%, and 46.6%, and at 50% tracing completeness they were 20.5%, 70.1%, and 66.4%. These threshold-based summaries provide an alternative view of the upper tail of future outbreak risk, highlighting

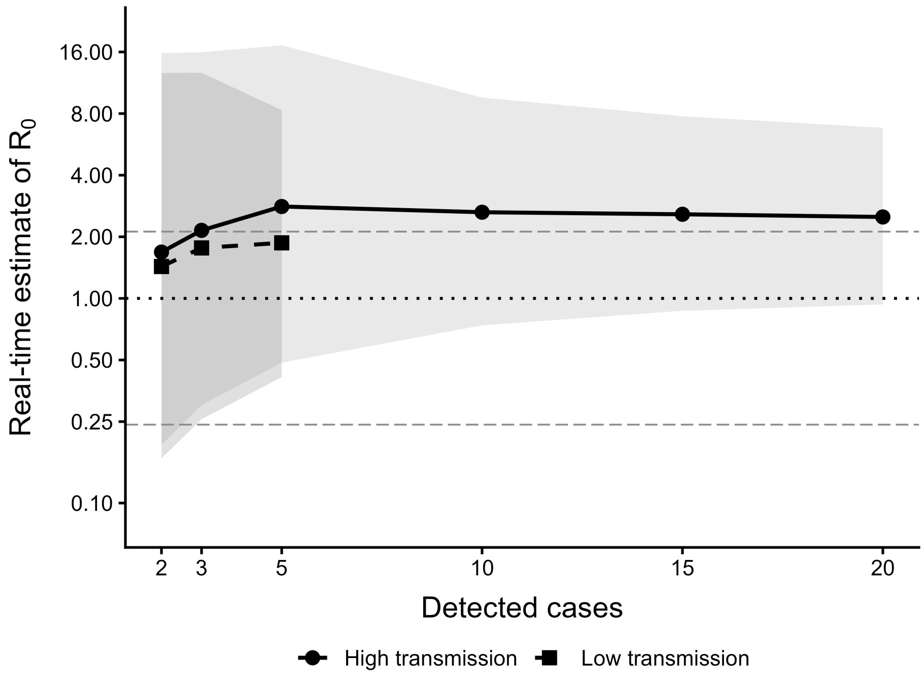


**Supplementary Figure S1: Real-time estimation of the reproduction number from accumulating detected cases under contrasting transmission scenarios.** Outbreaks were simulated under an Epuyén-calibrated higher-transmission scenario ($R = 2.12,\ k = 0.316$) and a pooled lower-transmission scenario ($R = 0.242,\ k = 0.400$). At successive detected-case thresholds, $R$ and $k$ were estimated using only detected transmission information available by that observation time. The likelihood accounted for incomplete case detection and right truncation arising because recently infected secondary cases may not yet have been detected. Points show the median posterior estimate of $R$ across simulations reaching each threshold, and shaded regions show the typical within-outbreak 95% credible interval, defined as the median lower and upper posterior limits across those simulations. The dotted horizontal line indicates $R = 1$; long-dashed horizontal lines indicate the generating reproduction numbers.

how modest tracing failures can sharply increase the probability of large downstream outbreaks.

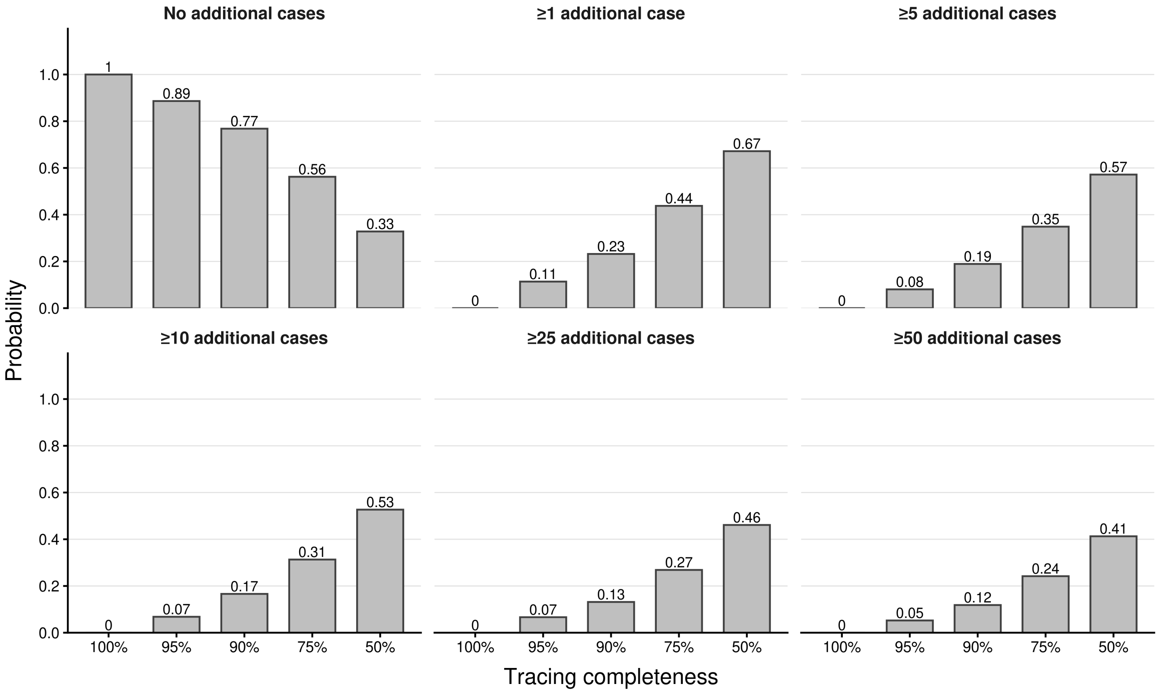


**Supplementary Figure S2: Threshold probabilities for additional infections after generation 2 under the Epuyén-calibrated higher-transmission scenario.** Simulations were conditioned on the uncontrolled potential transmission tree reaching generation 2 by the fixed response day. Bars show the estimated probabilities of no additional cases and of at least 5, 10, 25, or 50 additional infections after generation 2, stratified by tracing completeness.

#### Outbreak-size distributions

We examined simulated outbreak-size distributions to assess whether outcomes separated into distinct groupings and whether a natural breakpoint could be identified between self-limited and larger outbreaks. Specifically, we examined both total infections over the full simulated outbreak and additional infections after the current generation-2 response point, stratified by tracing completeness and by the generation at which tracing and quarantine began.

The simulated outbreak-size distributions were generally strongly right-skewed, with many simulations producing few infections and a smaller number producing substantially larger outbreaks. When tracing and quarantine began at generation 0 or 1, most simulations remained small under high tracing completeness. For example, at 95% tracing completeness, the 95th percentile of total infections was 5 when tracing began at generation 0 and 6 when tracing began at generation 1. The probability of at least 50 additional infections after generation 2 was correspondingly low, at 3.9% and 4.1%, respectively.

Delayed tracing and quarantine produced much heavier upper tails. At 95% tracing completeness, the 95th percentile of total infections increased to 383 when tracing began at generation 2, 978 when tracing began at generation 3, and 1,033 when tracing began at generation 4. The probability of at least 50 additional infections after generation 2 increased from 6.6% with tracing initiated at generation 2 to 14.1% with tracing initiated at generation 3 and 26.0% with tracing initiated at generation 4. Under the latest-start scenario, reducing tracing completeness from 100% to 50% increased the probability of at least 50 additional infections from 0.0% to 80.0%, and increased the 95th percentile of total infections from 200 to 4,040.

Although delayed-start scenarios showed a more apparent two-group pattern, with a concentration of smaller outbreaks and a broader upper-tail region of larger outbreaks, the location of the apparent valley between these regions varied across tracing-completeness and delayed-start assumptions. We therefore did not identify a single stable natural breakpoint that applied across scenarios. For this reason, we did not define large outbreaks using a single data-derived cutoff. Instead, we report multiple operationally interpretable threshold probabilities, including the probabilities of at least 5, 10, 25, and 50 additional infections after generation 2.

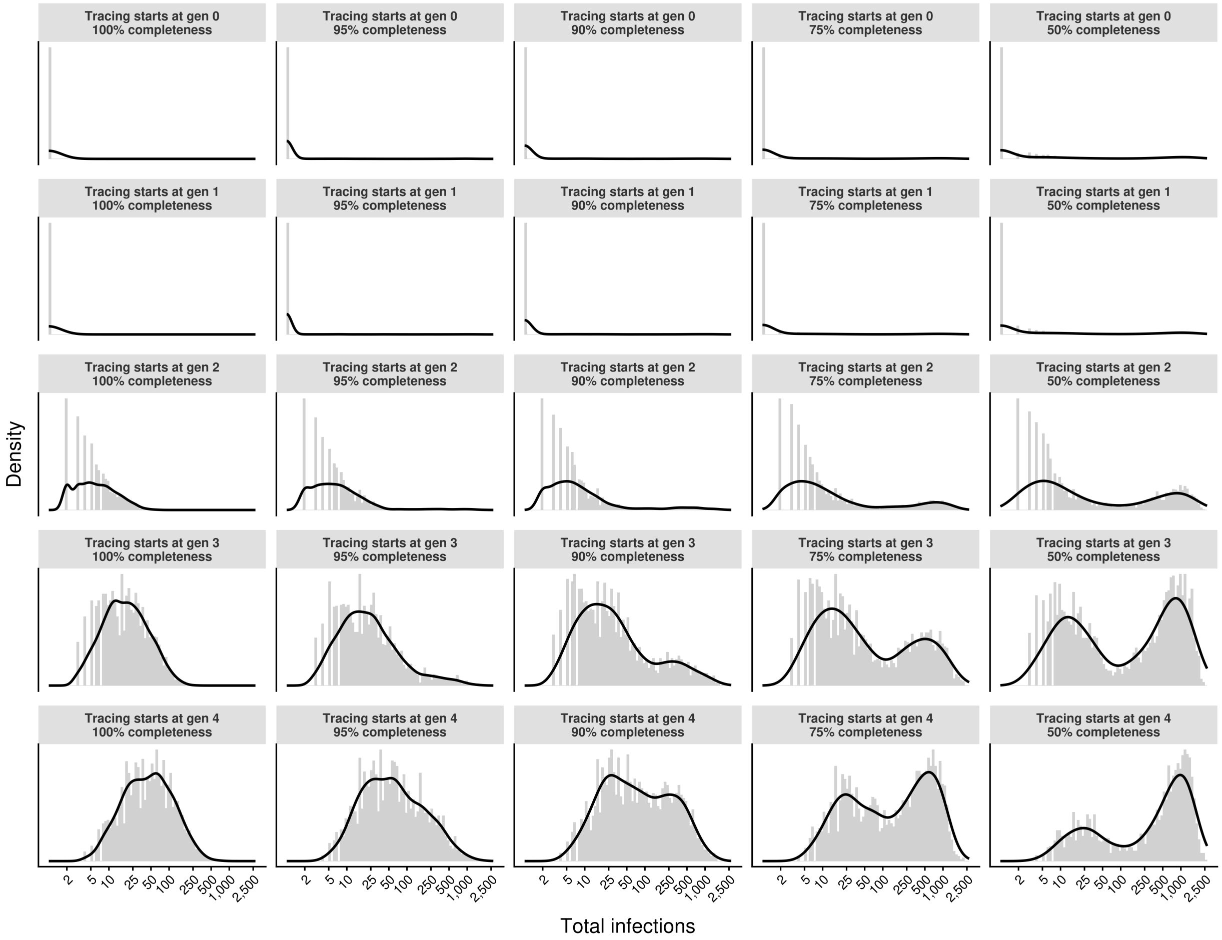


**Supplementary Figure S3: Outbreak-size distributions under generation-start tracing and quarantine sensitivity scenarios.** Each panel shows the simulated distribution of total retained infections among stochastic replicates that reached both the tracing-start generation and the current generation-2 response point. Columns correspond to tracing completeness and rows correspond to the generation at which tracing and quarantine began. Distributions are shown on a log-scaled $x$-axis to emphasize both self-limited outbreaks and the upper tail of larger outbreaks.
